# Subregional Functional Connectivity of the Precuneus as a Preclinical Biomarker in Alzheimer’s Disease

**DOI:** 10.1101/2025.04.15.25325852

**Authors:** Claudia Aponte, Antonio Jimenez-Marin, Malen Razkin, John Fredy Ochoa Gómez, Carlos Tobón, Asier Erramuzpe, Ibai Diez, David Aguillon-Niño, Jesus M. Cortes

## Abstract

**Background:** While the precuneus’ role in integrating diverse brain functions and its early involvement in Alzheimer’s Disease (AD) is well established, the differential impact of AD pathology on its subregions is poorly understood. This study aims to delineate the differential involvement and vulnerability of these subdivisions in the early stages of AD progression.

**Methods:** We conducted a resting-state functional connectivity analysis in 32 asymptomatic carriers of the PSEN1 E280A mutation for familial Alzheimer’s disease and compared them to 25 non-carrier familial controls. Seed-based functional connectivity analysis was applied to the precuneus and its subregions.

**Results:** Among carriers, the 7Am subregion exhibited the most pronounced statistical differences, consisting of increased connectivity with the entorhinal cortex, superior temporal gyrus, insula-operculum, dorsolateral prefrontal cortex, and somatosensory areas. The POS2 subregion further significantly decreased its connectivity with the anterior insula and dorsolateral prefrontal cortex. Higher MoCA scores correlated with increased within and between precuneus and frontoparietal network connectivity, alongside decreased connectivity between 7Pm, PCV, POS2, and the medial temporal lobe. Additionally, the 7m subregion displayed significantly higher connectivity with medial and dorsolateral prefrontal regions.

**Conclusions:** Our findings highlight the importance of subregional analysis in precuneus connectivity, uncovering patterns that do not exist when the precuneus is treated as a single region of interest—as is common in neuroimaging studies. Notably, even in preclinical stages of Alzheimer’s disease, early connectivity changes are evident, supporting their potential as biomarkers of disease progression. These results also point to the distinct involvement and vulnerability of precuneus subdivisions during the initial phases of AD.

## INTRODUCTION

The precuneus (Pc), a central hub within the posterior medial parietal lobe, is a critical integrative region implicated in diverse cognitive functions. Because of its high metabolic demands, the Pc is especially vulnerable to disease (1–7), with early signs of dysfunction in Alzheimer’s disease, exhibiting amyloid-β plaque and tau tangle accumulation earlier than other regions, leading to functional disconnection and atrophy, often years before clinical onset (8–13). The neurotoxicity of these proteins disrupts the functional connectivity of the PC (14,15), triggering a cascade of progressive disconnection across both functional and anatomical brain networks (16). Consequently, the resting-state network (RSN) connectivity patterns involving the Pc are altered. Among these networks, the Default Mode Network (DMN)—of which the Pc is a central hub— shows the most consistently reported connectivity disruptions when compared to healthy controls (6,16–20).

However, intriguing case studies of PSEN1 mutation carriers for Autosomal-Dominant Alzheimer’s Disease (ADAD) who remained cognitively resilient despite substantial amyloid and tau burden, revealed preserved Pc integrity, specifically the absence of tau accumulation and metabolic decline (21,22). This suggests a pivotal role for the Pc in mediating AD-related cognitive decline. Furthermore, studies employing Pc stimulation have shown promise in delaying AD progression (23). A deeper understanding of the functional connectivity patterns within Pc subregions, particularly their vulnerability during preclinical AD, is of maximum relevance for comprehension of early disease progression and inform the development of targeted neuromodulation strategies for AD.

A major complication for assessing connectivity of the Pc is its high heterogeneity, characterized by distinct functional subregions and variations in morphology. The anterior portion of the Pc mainly integrates the information from somatosensory areas, its posterior area integrates visual signals, and its medial region is involved in visuospatial integration, episodic memory, self-awareness and attention (24,25). Its medial portion varies in size in humans and does not exhibit a consistent morphological pattern (24). There also exist slightly morphological differences between hemispheres, the right Pc is larger than the left, with different shape and sulcal folding patterns variations between subjects (26).

To analyze the functional connectivity of Pc, several atlases have been developed to address the lack of homogeneity of cytological, anatomical and functional characteristics of this region. Some have cytoarchitectonic approaches (27–30), others are based on stereotaxic resonance imaging (31) and a combination of cytoarchitecture and tractography (32) and structural and functional connectivity (33–36) with a poor consensus on the number of regions the Pc is divided. Nonetheless, it has been proposed that the heterogeneity of structural connectivity across Pc subregions underlies the diversity of their functional connectivity profiles (25,34). Therefore, investigating each subregion independently is essential to advance our understanding of the functional role of the Pc, its involvement in higher-order cognitive processes, and its potential role in mediating the progression of neurodegenerative diseases such as AD.

Here, we hypothesized that specific Pc subfields would demonstrate enhanced biomarker sensitivity for early detection of AD compared to the whole Pc, and that these subfields would show varying degrees of vulnerability across disease stages. To do so, we focused on the functional connectivity of Pc and its sub-regions in asymptomatic carriers of PSEN1 E280A mutation compared to non-carriers. This population provides a valuable model as PSEN1 E280A carriers show early amyloid accumulation in the Pc (37) and altered connectivity during preclinical stages (38), mild cognitive impairment onset at ∼44 years, and dementia around 49 years (39). Notably, this study leverages data from the world’s largest familial aggregate of ADAD, located in Colombia, comprising over 6,000 family members and an estimated 1,200 PSEN1 E280A mutation carriers (40–43).

## METHODOLOGY

### Participants

A total of 32 asymptomatic carriers of the PSEN1 E280A mutation and 25 non-carriers—members of the same Colombian familial cluster with ADAD— were included in the study. Participants were matched by sex, age, and educational level, and all underwent neurological examinations, neuropsychological assessments, and brain magnetic resonance imaging. All participants were chosen from Neuroscience Group of Antioquia (GNA by its Spanish acronym) database, randomized 1:1 according to carrier versus non-carrier status, a total of 70 participants were selected but only 57 completed all procedures. Inclusion criteria were to belong to PSEN1 E280A mutation kindred and confirmed their condition of carrier before selection, cognitively asymptomatic with memory complaint scale <19 and without visual nor auditory disabilities documented in clinical history. Exclusion criteria was a mini-mental state examination (MMSE) score <26 in asymptomatic subjects, any significantly medical finding in the physical and neurological examination, history of traumatic brain injury, consumption of anticonvulsants, opiates, benzodiazepines or antidepressants drugs, consumption of psychoactive substances and any contraindication to undergo magnetic resonance imaging. All participants provided informed consent prior to study participation, in accordance with protocols approved by local institutional review boards. The study was conducted between January and September 2023. Participant recruitment, enrollment, clinical and neuropsychological evaluations, as well as magnetic resonance imaging acquisitions, were carried out at the University of Antioquia, Colombia.

### Neuropsychological and neurological examination

All participants were evaluated with the same neuropsychological tests consisting of Consortium to Establish a Registry for Alzheimer’s Disease (CERAD) adapted and validated to Colombian population (39,44), Trail Making Test -A part (TMT-A) (45), Rey-Osterrieth complex figure (46,47) copy and 5 minutes recall, FAS test (48), Raven’s Progressive Matrices part A (49), abbreviated Wisconsin Card Sorting Test (WCST) (50), memory impairment screen (MIS) (51), memory capacity test (MCT) (52), WAIS-III digit symbol Coding (53), Montreal Cognitive Assessment (MoCA) (54), INECO Frontal Screening (55), Functional Assessment Screening Tool (FAST) (56,57), Global Deterioration Scale (GDS) (58), Barthel Index for Activities of Daily Living (ADL)(59), Katz Index of Independence in Activities of Daily Living (60,61), The Lawton – Brody Instrumental Activities of Daily Living Scale (IADL) (62), Memory Complaints Scale and The Geriatric Depression Scale (GDS) (63).

Participants were excluded if they met diagnostic criteria for mild cognitive impairment or dementia, based on a comprehensive neuropsychological evaluation showing performance below 2 standard deviations on test scores and the clinical judgment of the evaluator. Additionally, all participants underwent a neurological assessment conducted by physicians from the GNA staff experienced in dementia evaluation, to identify any exclusion criteria. Inclusion and exclusion criteria were identical for both PSEN1 E280A mutation carriers and non-carriers; the only distinguishing factor between the two groups was the presence or absence of the mutation.

### MRI data acquisition

The MRI acquisition was performed on Philips Ingenia Elition X 3T scanner using a 32-channel head coil. A high-resolution 3D T1-weighted magnetization prepared rapid gradient-echo (MPRAGE) sequence was used with the following parameters: Field of view (FOV) = 256x256x211 mm, voxel size 1x1x1 mm (isotropic voxels), repetition time (TR) = 6200 ms, echo time (TE) = 2800 ms, flip angle 9.

Resting-state functional MRI (rs-fMRI) was acquired over a 10-minute scan with participants instructed to keep their eyes open, remain awake, and follow the instruction: “try not to think about anything specific.” Bi-temporal foam pads were used to minimize head motion. Imaging was performed using a single-shot echo planar imaging (EPI) sequence with the following parameters: field of view (FOV) = 256 × 256 × 180 mm, voxel size = 3 × 3 × 3 mm (isotropic), echo time (TE) = 27 ms, and repetition time (TR) = 2200 ms.

### MRI preprocessing

Pre-processing of images was performed with the Functional Connectivity Toolbox (CONN) (64), consisting of functional data realignment with correction for susceptibility distortion interactions, slice-timing correction, and co-registration with the T1-weighted anatomical image using a least-squares approach and a 6-parameter (rigid body) transformation. Resampling was performed using b-spline interpolation to correct for motion and magnetic susceptibility interactions. Direct segmentation and normalization to MNI space were subsequently applied. Participants were excluded if they exhibited excessive head motion, defined as frame-wise displacement (FD) >1 deg. of rotation or >1 mm of translation in any direction, based on the Artifact Detection Tools (ART) method (65). Additional exclusion criteria included global BOLD signal changes exceeding 2 standard deviations and more than 14 outlier volumes per session. First-level covariates included standard motion parameter time series and scrubbing regressors derived from ART to account for signal artifacts (66). Linear confound regression was performed to remove physiological and motion-related noise. This included five principal components each from cerebrospinal fluid and white matter (CompCor method), 12 motion regressors (six realignment parameters and their first-order derivatives), and additional nuisance regressors, including masked BOLD time series components, rest condition effects and their derivatives (2 factors), and linear trends (2 factors) within each functional run (67). Spatial smoothing was applied using a Gaussian kernel with a full width at half maximum (FWHM) of 6 mm (σ = 2.55). Finally, the data were temporally band-pass filtered to retain frequencies in the range of 0.008–0.09 Hz (68).

### Functional connectivity analysis of whole Pc and subregions

The whole Pc mask and its subregion masks were defined using coordinates from the Human Connectome Project Multi-Modal Parcellation atlas (33). This atlas delineates five subregions within the Pc: the precuneus visual area (PCV), parieto-occipital sulcus 2 (POS2), and three Brodmann area 7 subregions (7Am, 7m, and 7Pm), as illustrated in Figure 1. All subregions were defined bilaterally, and connectivity values were averaged across hemispheres for the analysis. In addition to subregional analyses, we included the whole Pc as a single region of interest, as a control for our study.

**Figure 1.**
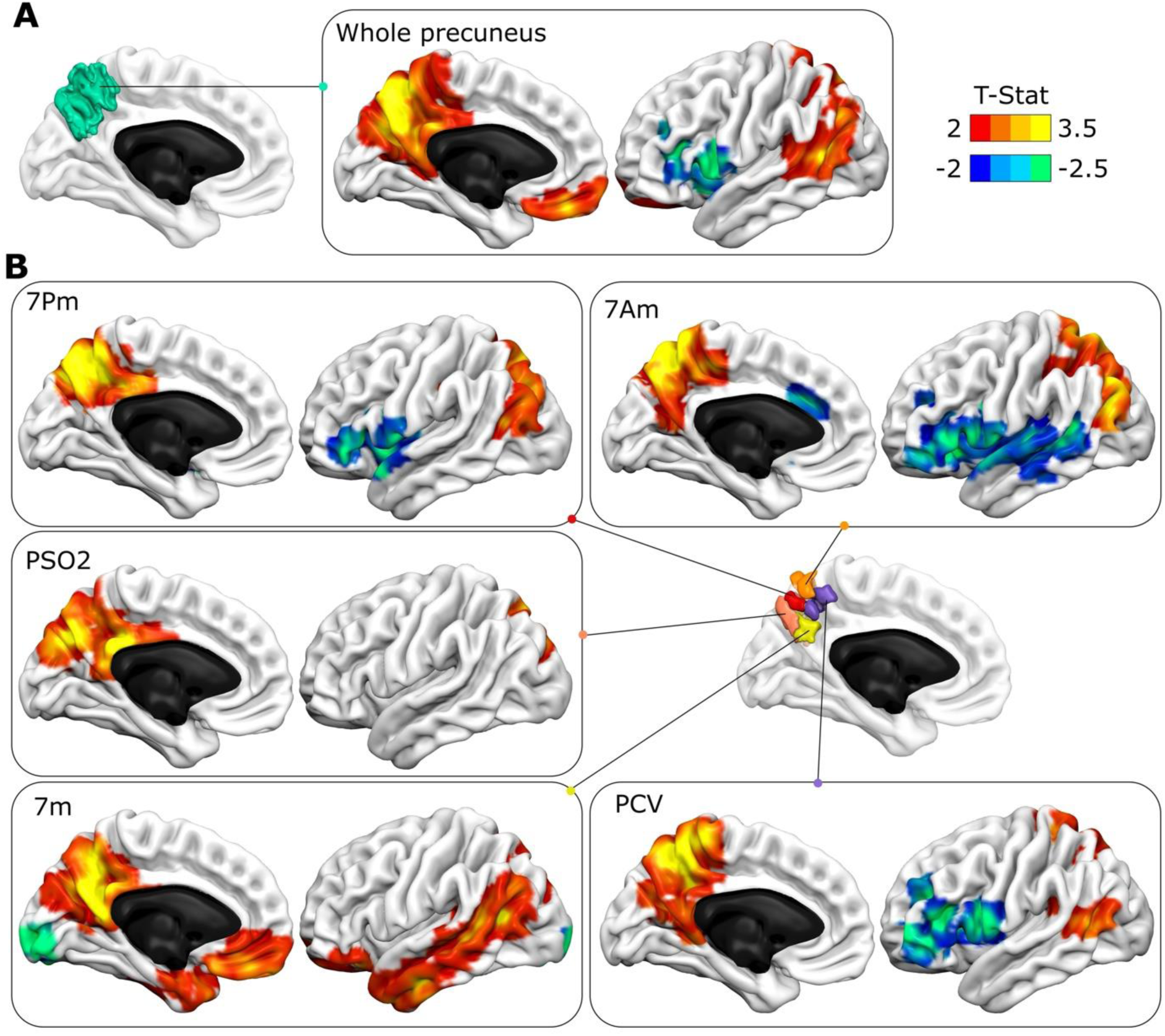
Seed-based connectivity (SBC) maps of the whole precuneus and its subregions. SBC maps were generated for the non-carrier group. Positive functional connectivity is indicated by a red-yellow color scale, while negative connectivity is shown in blue-green. **A:** Connectivity map for the whole precuneus, with the seed region highlighted in green. **B:** SBC maps for five precuneus subregions, illustrating distinct connectivity patterns compared to the whole precuneus. Abbreviations: 7Pm = Brodmann area 7 posteromedial; 7Am = BA7 anteromedial; 7m = BA7 medial; PCV = precuneus visual area; PSO2 = parieto-occipital sulcus 2.

Connectivity maps for each participant were generated for the five Pc subregions and the whole Pc using seed-based correlation (SBC) analysis. This approach involved computing the Pearson correlation between the BOLD time series of each seed region and all other voxels in the brain. The resulting individual connectivity maps were then used for group-level comparisons between carriers and non-carriers, with sex, age, and years of education included as covariates.

## STATISTICAL ANALYSIS

All statistical analyses were performed using the CONN toolbox (64), which provides validated pipelines for both image preprocessing and group-level neuroimaging analyses. SBC maps, represented the amount of functional connectivity between the seed and all other voxels in the brain, were calculated for the whole Pc and its subregions by using Fisher-transformed bivariate Pearson correlation coefficients derived from the BOLD time series. Group comparisons were conducted using the General Linear Model with Ordinary Least Squares estimation. Hypothesis testing was carried out using the Likelihood Ratio Test and Wilks’ Lambda distribution. Between-group differences were assessed with two-sample t-tests, while one-sample t-tests were used to evaluate average connectivity patterns within each group. Additionally, one-way ANCOVA models were applied to explore the interaction effects of covariates (sex, age, and years of education) on functional connectivity. Finally, cluster-level inferences were performed based on Gaussian Random Field theory. All reported results correspond to clusters that survived false discovery rate (FDR) correction for multiple comparisons at the cluster level. The anatomical localization of group differences was determined using region labels from the Harvard-Oxford atlas (69). For this study, only brain regions contributing to each significant cluster with at least 20% overlap were reported.

## RESULTS

Two groups were included in the study: 32 asymptomatic PSEN1 E280A mutation carriers (aC) and 25 non-carriers (nC). No fMRI datasets were excluded due to artifacts or outlier detection. Following neurological and neuropsychological assessments, none of the participants met diagnostic criteria for cognitive impairment (see Table S1). A significant group difference was observed in years of education, with the nC group having more years of schooling than the aC group (p < 0.001, 95% CI: 1.43–5.15). No significant differences were found in age or sex distribution between the groups. Additional demographic and clinical data are provided in Table 1.

**Table 1.** Demographic data of study participants.

| **Group**<br>**n= 32/25** | **Sex**<br>**(F/M)** | **Age**<br>**(mean/**<br>**SD)** | **Years of**<br>**school**<br>**(mean/SD**<br>**)** | **MMSE**<br>**score**<br>**(mean/S**<br>**D)** | **MoCA**<br>**score**<br>**(mean/S**<br>**D)** | **GDS**<br>**(mean/**<br>**SD)** |
| --- | --- | --- | --- | --- | --- | --- |
| aC | 21 F/11 | 35.29 | 10.35 | 27.9 | 24.1 | 1.22 |
|  | M | (5.44) | (2.80) | (1.86) | (3.95) | (0.491) |
| nC | 16 F/9 | 35.78 | 13.26 | 28.8 | 25.7 | 1 |
|  | M | (6.16) | (2.99) | (1.21) | (2.41) | (0.289) |

*aC: asymptomatic carrier; nC: non-carrier; F: female; M: male; SD: standard deviation; MMSE: Mini-mental state examination; MoCA: Montreal Cognitive Assessment; GDS: Global Deterioration Scale.*

To illustrate the differential capabilities of SBC maps using the whole Pc versus its subregions as seeds, we conducted an initial analysis in the group of aC, as presented in Figure 1. Specifically, SBC maps were generated using the whole Pc as a seed, along with the subregions 7m, 7Am, 7Pm, PCV, and POS2. In these maps, positive functional connectivity is depicted using a red-yellow color scale, while negative connectivity is shown in blue-green. A comprehensive anatomical description of the significant clusters identified for each seed (whole Pc and subregions) is provided in Tables S2–S7.

### Whole precuneus connectivity differences between Cognitively Unimpaired Mutation Carriers and Noncarriers

SBC analysis using the whole Pc revealed three clusters with significant group differences (Figure 2). Note that in the figure, the regions are color-coded to distinguish connectivity differences which are specific for whole Pc (clusters in blue) and those shared with subregions (cluster in yellow). Specifically, aC>nC connectivity was found in inter-DMN cluster 1 (p-FDR < 0.001; max t = 4.88), including right occipito-temporal regions, and in cluster 3 (p-FDR < 0.05; max t = 3.82), with no dominant anatomical overlap. aC<nC connectivity was observed in intra-DMN cluster 2 (p-FDR < 0.001; max t = 4.44), involving the medial frontal cortex. Full cluster details are given in Table S8.

**Figure 2:**
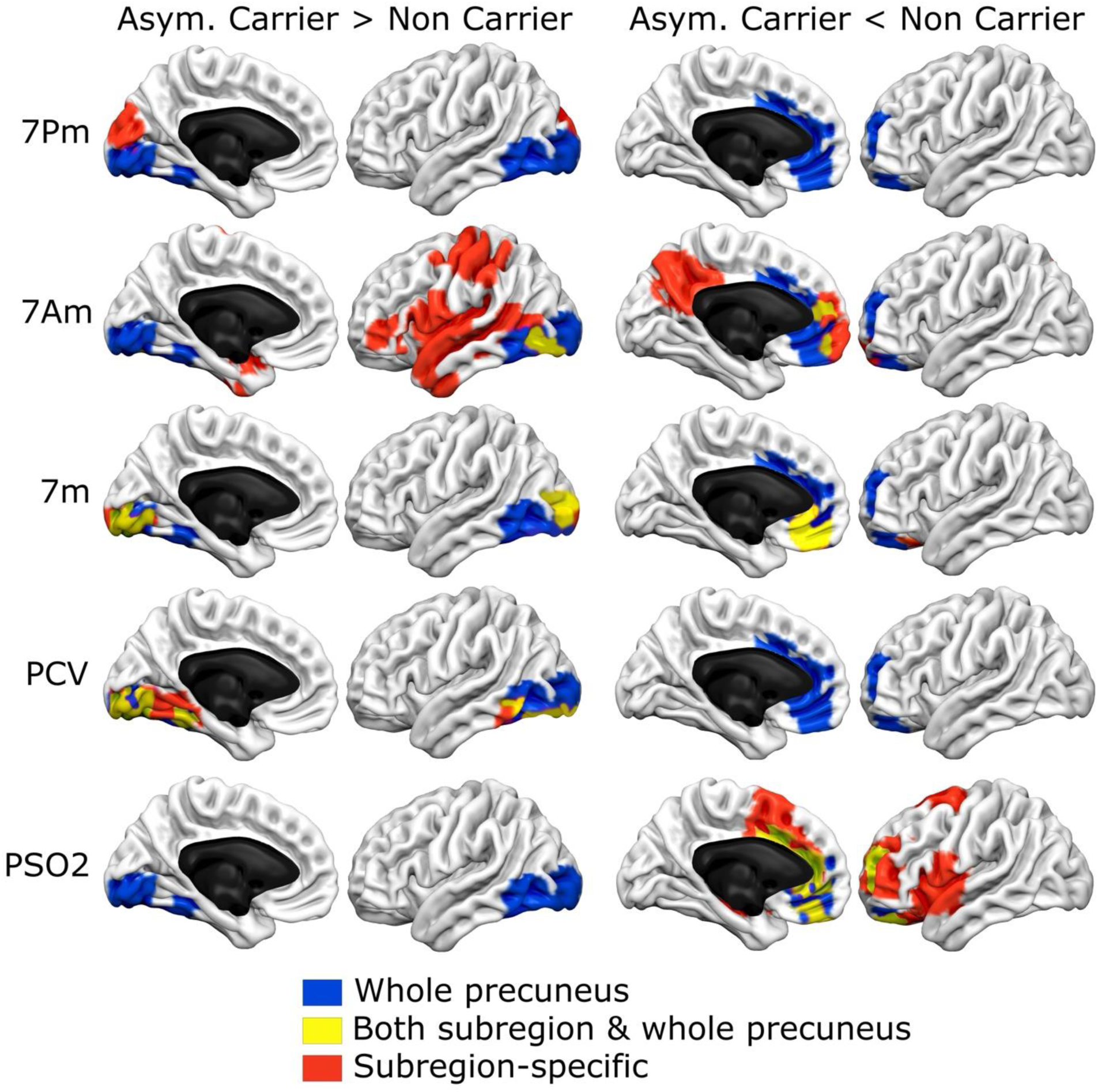
Differential impact of using the whole precuneus vs. its subregions in detecting connectivity changes in Alzheimer’s disease. This figure illustrates the advantage of subdividing the precuneus when computing connectivity differences between asymptomatic PSEN1 E280A mutation carriers (aC) and non-carriers (nC). Connectivity differences (aC > nC: hyperconnectivity, left column; aC < nC: hypoconnectivity, right column) are color-coded in the following manner: Blue: Differences identified using the whole precuneus as the seed. Yellow: Differences observed in each subregion that also appear when using the whole precuneus. Red: Differences detected only when using individual subregions as seeds. Notably, the overlapping yellow patterns across all subregions closely resemble those found using the whole precuneus (blue), validating its general sensitivity. However, the red areas reveal additional, unique information captured only through subregional analysis, highlighting the differential value of this approach. Abbreviations: 7Pm = Brodmann area 7 posteromedial; 7Am = BA7 anteromedial; 7m = BA7 medial; PCV = precuneus visual area; PSO2 = parieto-occipital sulcus 2.

### Subregion connectivity differences between Cognitively Unimpaired Mutation Carriers and Noncarriers

<u>7m subregion:</u> SBC revealed two clusters with significant group differences (Figure 2). aC<nC connectivity was observed in cluster 1 (p-FDR < 0.05; max t = 4.58), involving the medial frontal cortex. aC>nC connectivity was found in cluster 2 (p-FDR < 0.05; max t = 3.30), located in the right occipital pole. Full cluster details are given in Table S9.

### 7Am subregion

Nine clusters showed significant group differences (Figure 2). aC>nC connectivity was found in clusters 1 (p-FDR < 0.001; max t = 3.86; left opercular and temporal regions), 3 (p-FDR < 0.001; max t = 3.92; right middle temporal gyrus), 5 (p-FDR < 0.05; max t = 4.01; right insula and inferior temporal gyrus), 6 (p-FDR < 0.05; max t = 3.37), and 9 (p-FDR < 0.05; max t = 3.61; left lingual gyrus). aC<nC connectivity was observed in clusters 2 (p-FDR < 0.001; max t = 4.15; Pc and posterior cingulate), 4 (p-FDR < 0.05; max t = 4.73), 7 (p-FDR < 0.05; max t = 3.57), and 8 (p-FDR < 0.05; max t = 3.64). Full cluster details are given in Table S10.

### 7Pm subregion

One cluster showed significant group differences (p-FDR < 0.05; max t = 3.31), with aC>nC connectivity to the left lingual gyrus (Figure 2). Full cluster details are given in Table S11.

### PCV subregion

One cluster showed significant group differences (p-FDR < 0.05; max t = 3.96), with aC>nC connectivity to the right lingual gyrus, occipital fusiform gyrus, and inferior temporal gyrus (temporo-occipital part) (Figure 2). Full cluster details are given in Table S12.

### POS2 subregion

Two clusters showed significant group differences (Figure 2), both with aC<nC connectivity. Cluster 1 (p-FDR < 0.001; max t = 4.36) involved the anterior cingulate gyrus; cluster 2 (p-FDR < 0.05; max t = 3.67) included the right insular cortex, central opercular cortex, and planum polare. Full cluster details are given in Table S13.

Note that in Figure 2, regarding the connectivity differences in the SBC maps of the different Pc subregions, the clusters being referred to are those highlighted in red (specific to each subregion) and yellow (shared differences with whole Pc).

### Whole precuneus connectivity association with cognitive functioning for Asymptomatic Mutation Carriers

Association between SBC from whole Pc and MOCA’s scores revealed three significant clusters (Figure 3). Note that in the figure, the regions are color-coded to distinguish connectivity differences which are specific for whole Pc (clusters in blue) and those shared with subregions (cluster in yellow). Specifically, it was found a positive association in cluster 1 (p-FDR < 0.001; max t = 7.35, intra-DMN; precuneus and posterior cingulate cortex), 3 (p-FDR < 0.05; max t = 3.05; medial frontal cortex). A negative association was found in cluster 2 (p-FDR < 0.005; min t = -4.10; right occipito-temporal). Full cluster details are given in Table S14.

**Figure 3:**
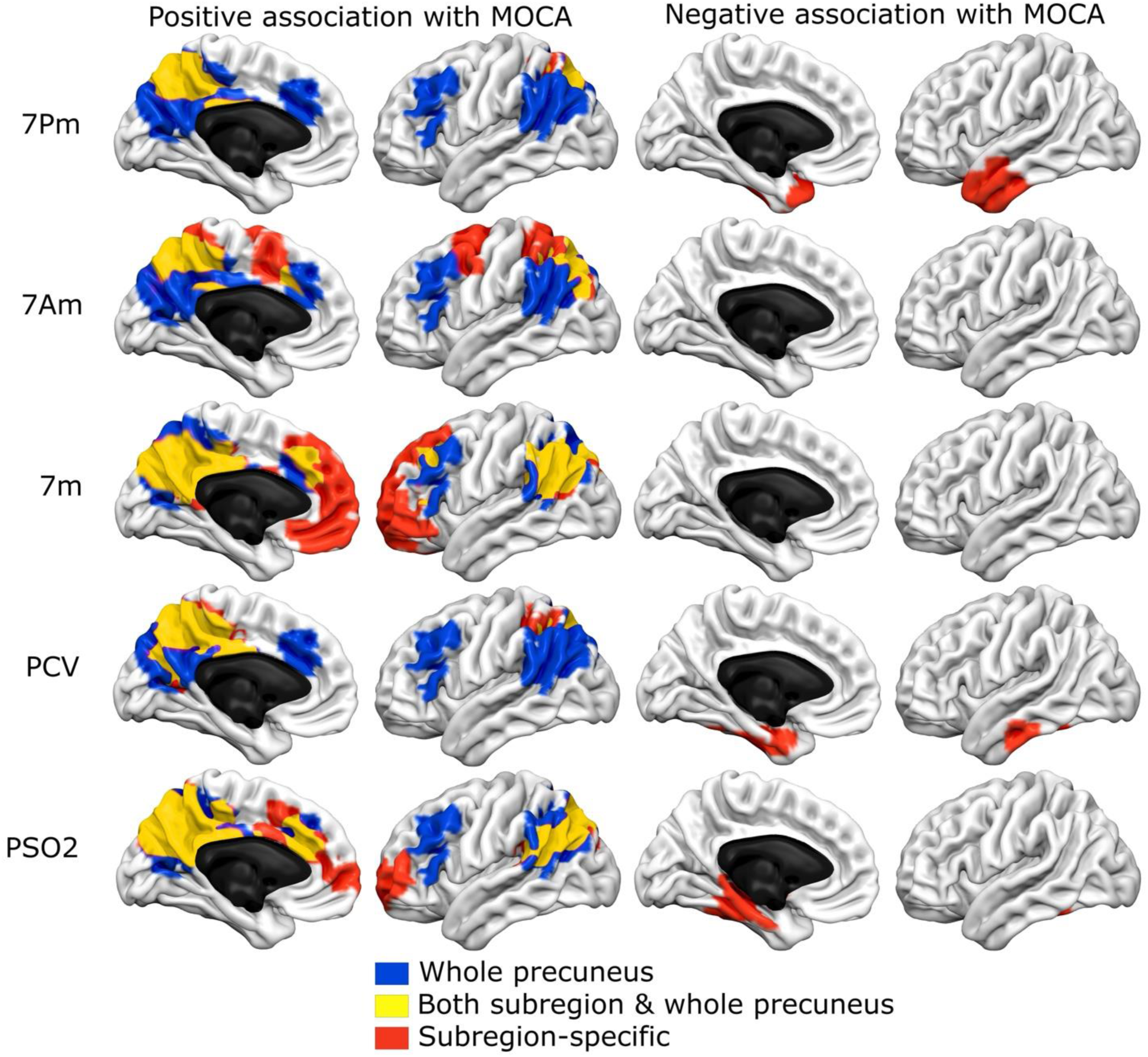
Association between global cognition (MOCA) and connectivity patterns of the precuneus and its subregions. The statistically significant association between global cognitive performance (as measured by the MoCA test) and SBC maps of the precuneus and its subregions was assessed. Color coding follows the same scheme as in figure 3. Blue: Associations detected using the whole precuneus as the seed. Yellow: Overlapping associations observed in both the whole precuneus and individual subregions. Red: Associations identified only when using subregions as seeds. The yellow overlap across all subregions closely resembles the blue pattern, indicating that subregional analysis retains the information captured by the whole precuneus. Importantly, additional associations (in red) emerge only at the subregional level, demonstrating the differential value of a more fine-grained approach. Abbreviations: 7Pm = Brodmann area 7 posteromedial; 7Am = BA7 anteromedial; 7m = BA7 medial; PCV = precuneus visual area; PSO2 = parieto-occipital sulcus 2.

### Subregion connectivity association with cognitive functioning for Asymptomatic Mutation Carriers

#### 7Pm subregion

Four significant clusters (Figure 3). A positive association was found in cluster 1 (p-FDR < 0.001; max t = 7.35; intra-DMN, precuneus and posterior cingulate cortex). A negative association was found in cluster 2 (p-FDR < 0.05; min t = -4.45), 3 (p-FDR < 0.05; min t = -3.40), and 4 (p-FDR < 0.05; min t = -3.80), all of them related to left and right temporal regions. Full cluster details are given in Table S15.

#### 7Am subregion

Two significant clusters (Figure 3). Cluster 1 (p-FDR < 0.001; max t = 8.40; precuneus and lateral occipital cortex), 2 (p-FDR < 0.005; max t = 3.40; superior frontal gyrus). Full cluster details are given in Table S16.

#### 7m subregion

Six significant clusters (Figure 3). Cluster 1 (p-FDR < 0.001; max t = 6.05; intra-DMN, precuneus and posterior cingulate cortex), 2 (p-FDR < 0.001; max t = 4.60; frontal pole and paracingulate gyrus), 3 (p-FDR < 0.001; max t = 5.60; right lateral occipital cortex and angular gyrus), 5 (p-FDR < 0.005; max t = 4.70; left lateral occipital cortex and angular gyrus), 4 (p-FDR < 0.001; max t = 5.00; superior frontal and paracingulate gyri), and 6 (p-FDR < 0.05; max t = 4.40; middle frontal gyrus). Full cluster details are given in Table S17.

### PCV subregion

Two significant clusters (Figure 3). A positive association was found in cluster 1 (p-FDR < 0.001; max t = 8.50; intra-DMN, precuneus and posterior cingulate cortex). A negative association was found in cluster 2 (p-FDR < 0.05; min t = -3.75; brain stem, inferior temporal gyrus and cerebellum crus 1). Full cluster details are given in Table S18.

### POS2 subregion

Five significant clusters (Figure 3). A positive association was found in cluster 1 (p-FDR < 0.001; max t = 7.70; intra-DMN, precuneus and posterior cingulate cortex), 3 (p-FDR < 0.005; max t = 3.55; right frontal pole and right paracingulate gyrus), and 4 (p-FDR < 0.05; max t = 5.70; left frontal pole). A negative association was found in cluster 2 and 5 (p-FDR < 0.005; min t = -4.30; p-FDR < 0.05; min t = -3.65), both related to left and right hippocampus and parahippocampal gyri. Full cluster details are given in Table S19.

Note that in Figure 3, regarding the connectivity differences in the SBC maps of the different Pc subregions, the clusters being referred to are those highlighted in red (specific to each subregion) and yellow (shared differences with whole Pc).

For other cognitive tests, such as CERAD’s List of Words, and MCT (Figure S1), see complete description of significant clusters in Tables S20-S31.

## DISCUSSION

This study leverages a globally unique cohort of cognitively unimpaired carriers of the PSEN1 E280A mutation—considered one of the most robust preclinical models of autosomal dominant Alzheimer’s disease (ADAD)—to investigate early functional connectivity alterations within the precuneus (Pc). Building on converging evidence that the Pc is not only among the earliest regions affected by AD pathology but also a critical hub in individuals exhibiting cognitive resilience, we hypothesized that analyzing its subregions independently could reveal differential vulnerability patterns obscured when treating the Pc as a single region of interest. Our results strongly support this hypothesis, demonstrating that subregional analysis provides more nuanced and spatially specific insights into early functional changes associated with AD, which may be essential for identifying early biomarkers and therapeutic targets during the preclinical stage of the disease.

We compared cognitively unimpaired individuals from the same familial background, focusing on asymptomatic carriers (aC) of the PSEN1 E280A mutation and non-carriers (nC). aC exhibited increased functional connectivity with regions including the right occipital cortex, bilateral temporal cortex, bilateral lingual gyri, left frontal cortex, and right insular cortex—areas reflecting inter-network connectivity between the DMN and other resting-state networks. Conversely, aC showed reduced connectivity within regions primarily associated with intra-DMN connectivity, such as the cingulate gyrus, medial frontal cortex, central opercular cortex, right insular cortex, right planum polare, and the Pc itself. These patterns are consistent with previous findings in PSEN1 E280A aC (38), indicating less segregated and integrated functional connectivity in the Pc. Such alterations are thought to result from the neurotoxicity of amyloid and tau proteins (70–72), which disrupt neural network communication before cognitive deficits emerge (73). The Pc is known to accumulate amyloid (8,74,75) and tau (75,76) early in both sporadic and ADAD (37,77–80), and this accumulation has been linked to hypometabolism (80–82) and altered task-based and resting-state connectivity (14,37,38,83,84) even during preclinical stages. These findings suggest that the neurotoxic effects of amyloid and tau impair metabolic and functional network integrity in the Pc years before symptom onset.

Despite both groups performing within the normal range on cognitive tests, significant differences in SBC maps may support the hypothesis that functional reorganization within and beyond the DMN acts as a mechanism of network resilience in ADAD (37,38,70,73,85), enabling the redistribution of information processing to maintain cognitive performance. This is consistent with previous findings showing that individuals carrying the PSEN1 E280A mutation—at an average age of 35—already present with elevated tau deposition in the medial and lateral temporal cortices, as well as in the Pc (38). These early molecular changes may contribute to functional disconnection within these networks. Notably, while hippocampal atrophy can already be detected at this stage, cortical atrophy remains absent, suggesting that functional disruptions precede measurable structural degeneration and may serve as early indicators of impending cognitive decline.

Among the subregions of the Pc, 7Am exhibited the most pronounced differences in functional connectivity between aC and non-carriers, as well as the most diverse connectivity profile. In aC, this region showed increased functional connectivity with the left and right lateral occipital cortex, right superior parietal lobule, left temporal fusiform cortex, and left parahippocampal gyrus. Given the association of 7Am with episodic memory retrieval, self-referential processing, and consciousness (86), the observed increase in connectivity may reflect a compensatory reorganization of information flow. This reorganization could support enhanced cognitive integration and associative processes, potentially offsetting the reduced connectivity observed in other posterior brain networks. Such mechanisms may underline the preserved cognitive performance in aC despite early functional connectivity changes.

In the case of the POS2 subregion, aC showed reduced functional connectivity, particularly with the insular cortex, anterior cingulate cortex, and opercular cortex. POS2 has been implicated in multitasking and visuospatial processing, especially in the identification of complex visual stimuli for subsequent integration into higher-order cognitive functions (87). The observed hypoconnectivity in aC may reflect a diminished capacity to process and integrate complex visual information, potentially impacting early stages of perceptual-cognitive integration. This interpretation aligns with previous behavioral findings using neurophysiological memory-binding paradigms, which revealed impairments in visual short-term memory binding in asymptomatic E280A carriers. Specifically, these individuals exhibited difficulties in maintaining unified object representations in visual short-term memory (88), suggesting early functional disruption in posterior Pc networks may contribute to subtle cognitive vulnerabilities despite preserved overall performance.

Across subregions, POS2 functional connectivity is primarily restricted to the posterior portion of the DMN. In contrast, PCV and 7m exhibit positive correlations with both lateral and anterior regions of the DMN, anatomically involving the temporal and frontal lobes. Notably, in aC, 7m and PCV show a lack of anticorrelations with the DMN, suggesting altered network dynamics. These findings highlight the distinct roles of Pc subregions in DMN connectivity: PCV and 7m may facilitate the distribution of information toward anterior brain regions through interactions with other networks, while other Pc subregions appear to sustain information flow within posterior association areas, potentially serving as core hubs for DMN integration. This pattern may indicate that, in the presence of emerging deficits in complex feature identification, enhanced cognitive association is required to support decision-making processes, compensating for early disruptions in posterior network function.

Interestingly, in this study, mutation carrier individuals who exhibited increased Pc connectivity with lateral frontoparietal regions—potentially compensating for medial temporal disconnection— also demonstrated higher MoCA scores (demonstrated normal cognition) (Figure 3). This finding suggests that Pc connectivity may support compensatory mechanisms that help preserve cognitive function despite underlying dysconnectivity caused by pathological protein accumulation. Prior studies have shown that enhanced connectivity between DMN structures and frontoparietal hubs is associated with delayed cognitive decline in AD (89–92). Thus, the Pc may serve a dual role across the AD continuum, contributing both to network vulnerability and to compensatory responses, with distinct subregions engaging in these processes differently.

This study has several limitations. One is the relatively small sample size; however, ADAD populations are rare, and not all members of the family cluster met the inclusion criteria or were willing to participate. Despite this, our findings are consistent with previous studies in the same PSEN1 E280A population, which have reported early alterations in the Pc. Specifically, prior research has shown that the earliest detectable changes in the DMN among carriers include Pc atrophy measured via volumetry (81,93), reduced glucose metabolism (81), and functional connectivity alterations during memory tasks and at rest (37,83,94,95). Another limitation is that the control group had a higher level of education, which could be a confounding factor. However, this variable was included as a regressor in the analysis, thus minimizing its effects on the results.

Despite these limitations, our study represents an important first step in exploring functional connectivity differences in preclinical stages of the AD continuum using resting-state fMRI (rs-fMRI) and SBC analysis. This approach successfully identified connectivity alterations between populations of interest and is the first of its kind applied to this cohort. More advanced connectivity methods, such as dynamic functional connectivity (dFC), could be employed in future studies to further characterize network dynamics in this population. Our findings align with previous reports and support the hypothesis that these connectivity changes may be linked to early amyloid accumulation in PSEN1 E280A mutation carriers (37,83,89). Taken together, these results highlight the potential value of examining precuneus and DMN connectivity as a neurophysiological marker for early detection and monitoring of AD progression.

To conclude, to the best of our knowledge, this is the first study in AD to analyze functional connectivity of the Pc by subdividing it into distinct subregions. As a result, direct comparisons with previous studies that examined Pc connectivity as a whole or in relation to other brain networks are limited. Our findings demonstrate that analyzing the Pc at a subregional level reveals connectivity differences that are not detectable when using the entire Pc or other subregions as a single seed. This underscores the importance of a more granular approach to functional connectivity analyses in preclinical AD.

## Data Availability

Data would be available upon reasonable request

## ACKNOWLEDGMENTS

The authors would like to thank the Ministerio de Ciencia, Tecnología E Innovación – Minciencias for its financial support to the project “Identificación de Biomarcadores Preclínicos en Enfermedad de Alzheimer a través de un Seguimiento Longitudinal de la Actividad Eléctrica Cerebral en Poblaciones con Riesgo Genético”, identified with the code 111577757635 contract 772-2017. JMC acknowledges financial support from Ikerbasque: The Basque Foundation for Science, and from Spanish Ministry of Science (PID2023-148012OB-I00), Spanish Ministry of Health (PI22/01118), Basque Ministry of Health (2023111002 & 2022111031).

## Author Contributions

Data curation: CA, AJM, MR. Methodology: CA, AE, ID, JMC. Investigation: CA, JFOG, CT, DAN, JMC. Data Analysis: CA, AJM, MR, ID. Supervision: JFOG, CT, JMC. Writing-original draft: CA. Writing-review and editing: All authors

