## Supplementary material for "Subregional Functional Connectivity of the Precuneus as a Preclinical Biomarker in Alzheimer’s Disease"

### Figure S1:

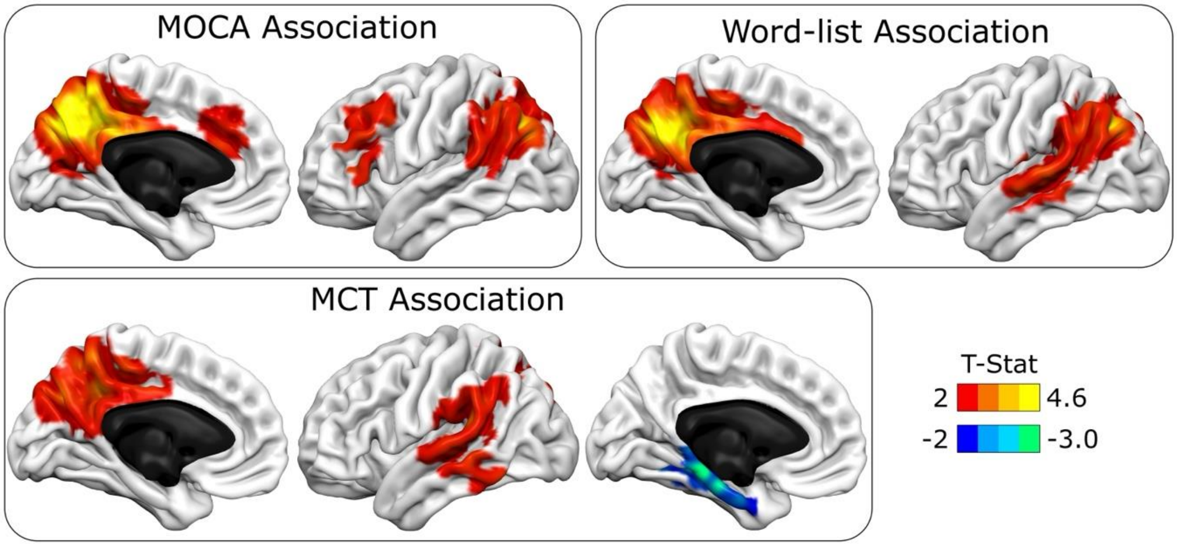


**Figure S1: Associations between whole precuneus connectivity and cognitive performance across different tests.** While Figure 3 shows the association between whole precuneus seed-based connectivity (SBC) and MOCA scores for simplicity, additional significant associations were also found with other cognitive measures. This figure shows SBC results using the whole precuneus as the seed, highlighting associations with word-list recall and the Memory Capacity Test (MCT), both of which display consistent connectivity patterns like those observed for MOCA. Consistent results were also observed when using precuneus subregions as seeds in the SBC analysis. Abbreviations: MOCA = Montreal Cognitive Assessment; MCT = Memory Capacity Test.

### Table S1. Neuropsychological test differences between asymptomatic carrier (aC) and non-carriers (nC).

| Group n= 32/25 | INECO_FS (mean/SD) | MIS_score (mean/SD) | List_words_ recall (mean/SD) | MCT_recall (mean/SD) | Digit_Simbol (mean/SD) | WSCT_cate gories (mean/SD) | WSCT_concep_ answ (mean/SD) | Memory_co mplains (mean/SD) |
|---|---|---|---|---|---|---|---|---|
| aC | 18.3 (6.68) | 6.34 (2.09) | 20.2 (5.21) | 20.2 (7.89) | 50.2 (22.1) | 2.88 (1.77) | 21.5 (11.1) | 7.78 (5.49) |
| nC | 21.3 (5.36) | 7.2 (1.66) | 21.7 (5.34) | 22.4 (6.48) | 64.9 (21.0) | 4 (1.94) | 27.8 (10.9) | 5.18 (3.77) |

*aC: asymptomatic carrier; nC: non-carrier; SD: standard deviation; INECO_FS: INECO Frontal Screening; MIS: Memory Impairment Screening; List_words_recall: late recall of the list of words in the CERAD test; MCT_recall: Free recall of the two list of words in the Memory Capacity Test; WSCT_categories: number of categories in the Wisconsin Sorting Card Test; WSCT_concep_answ: conceptual answers in the Wisconsin Sorting Card Test; Memory_complains: Memory Complains Test.*

**Table S2. SBC maps in asymptomatic carriers using the whole Precuneus as the seed.** The table reports the MNI coordinates of peak voxels for each cluster showing significant group differences, along with cluster size (in voxels) and p-values corrected for multiple comparisons (both at the cluster level and at the peak voxel level).

| Cluster (x,y,z) | Size (vox) | size p-FWE | size p-FDR | size p-unc | peak p-FWE | peak p-unc |
|---|---|---|---|---|---|---|
| -10 -58 +48 | 10531 | 0.000000 | 0.000000 | 0.000000 | 0.029820 | 0.000000 |
| +50 +14 +10 | 2668 | 0.000160 | 0.000065 | 0.000001 | 0.931835 | 0.000060 |
| +08 +48 -30 | 1048 | 0.109449 | 0.031143 | 0.000492 | 0.999159 | 0.000204 |
| +14 -16 -02 | 933 | 0.184992 | 0.034682 | 0.000868 | 0.999759 | 0.000252 |
| -38 -22 -22 | 923 | 0.193575 | 0.034682 | 0.000913 | 0.999989 | 0.000381 |

**Cluster -10 -58 +48**:
4024 voxels (38%) covering 72% of atlas.Precuneous (Precuneous Cortex)
1017 voxels (10%) covering 42% of atlas.PC (Cingulate Gyrus, posterior division)
839 voxels (8%) covering 17% of atlas.sLOC r (Lateral Occipital Cortex, superior division Right)
371 voxels (4%) covering 58% of atlas.Cuneal r (Cuneal Cortex Right)
367 voxels (3%) covering 7% of atlas.sLOC l (Lateral Occipital Cortex, superior division Left)
263 voxels (2%) covering 51% of atlas.Cuneal l (Cuneal Cortex Left)
217 voxels (2%) covering 19% of atlas.toMTG r (Middle Temporal Gyrus, temporooccipital part Right)
208 voxels (2%) covering 14% of atlas.AG r (Angular Gyrus Right)
154 voxels (1%) covering 4% of atlas.PostCG l (Postcentral Gyrus Left)
152 voxels (1%) covering 7% of atlas.iLOC r (Lateral Occipital Cortex, inferior division Right)
137 voxels (1%) covering 4% of atlas.PostCG r (Postcentral Gyrus Right)
109 voxels (1%) covering 9% of atlas.pSMG r (Supramarginal Gyrus, posterior division Right)
95 voxels (1%) covering 6% of atlas.SPL r (Superior Parietal Lobule Right)
53 voxels (1%) covering 8% of atlas.ICC l (Intracalcarine Cortex Left)
41 voxels (0%) covering 1% of atlas.PreCG l (Precentral Gyrus Left)
40 voxels (0%) covering 28% of atlas.SCC r (Supracalcarine Cortex Right)
27 voxels (0%) covering 2% of atlas.SPL l (Superior Parietal Lobule Left)
18 voxels (0%) covering 25% of atlas.SCC l (Supracalcarine Cortex Left)
10 voxels (0%) covering 1% of atlas.ICC r (Intracalcarine Cortex Right)
8 voxels (0%) covering 0% of atlas.PreCG r (Precentral Gyrus Right)
3 voxels (0%) covering 0% of atlas.Ver45 (Vermis 4 5)
1 voxels (0%) covering 0% of atlas.LG r (Lingual Gyrus Right)
2377 voxels (23%) covering 1% of atlas.not-labeled

**Cluster +50 +14 +10**:
487 voxels (18%) covering 36% of atlas.IC r (Insular Cortex Right)
250 voxels (9%) covering 80% of atlas.FO r (Frontal Operculum Cortex Right)
185 voxels (7%) covering 27% of atlas.IFG oper r (Inferior Frontal Gyrus, pars opercularis Right)
183 voxels (7%) covering 23% of atlas.Putamen r
117 voxels (4%) covering 13% of atlas.CO r (Central Opercular Cortex Right)
77 voxels (3%) covering 1% of atlas.FP r (Frontal Pole Right)
53 voxels (2%) covering 10% of atlas.IFG tri r (Inferior Frontal Gyrus, pars triangularis Right)
49 voxels (2%) covering 58% of atlas.Accumbens r
43 voxels (2%) covering 2% of atlas.AC (Cingulate Gyrus, anterior division)
42 voxels (2%) covering 1% of atlas.PreCG r (Precentral Gyrus Right)
34 voxels (1%) covering 13% of atlas.Pallidum r
33 voxels (1%) covering 2% of atlas.FOrb r (Frontal Orbital Cortex Right)
31 voxels (1%) covering 8% of atlas.PP r (Planum Polare Right)
25 voxels (1%) covering 1% of atlas.TP r (Temporal Pole Right)
7 voxels (0%) covering 0% of atlas.MidFG r (Middle Frontal Gyrus Right)

4 voxels (0%) covering 1% of atlas.Caudate r
3 voxels (0%) covering 1% of atlas.Amygdala r
1 voxels (0%) covering 0% of atlas.SubCalC (Subcallosal Cortex)
1044 voxels (39%) covering 0% of atlas.not-labeled

**Cluster +8 +48 -30**:
303 voxels (29%) covering 31% of atlas.MedFC (Frontal Medial Cortex)
234 voxels (22%) covering 3% of atlas.FP l (Frontal Pole Left)
163 voxels (16%) covering 2% of atlas.FP r (Frontal Pole Right)
71 voxels (7%) covering 6% of atlas.SubCalC (Subcallosal Cortex)
6 voxels (1%) covering 0% of atlas.FOrb l (Frontal Orbital Cortex Left)
271 voxels (26%) covering 0% of atlas.not-labeled

**Cluster +14 -16 -2**:
271 voxels (29%) covering 20% of atlas.Thalamus l
232 voxels (25%) covering 18% of atlas.Thalamus r
40 voxels (4%) covering 13% of atlas.Pallidum l
15 voxels (2%) covering 3% of atlas.Caudate l
8 voxels (1%) covering 1% of atlas.Hippocampus r
7 voxels (1%) covering 1% of atlas.Caudate r
2 voxels (0%) covering 1% of atlas.Pallidum r
1 voxels (0%) covering 0% of atlas.Amygdala r
357 voxels (38%) covering 0% of atlas.not-labeled

**Cluster -38 -22 -22**:
206 voxels (22%) covering 24% of atlas.pTFusC l (Temporal Fusiform Cortex, posterior division Left)
150 voxels (16%) covering 6% of atlas.TP l (Temporal Pole Left)
68 voxels (7%) covering 15% of atlas.aMTG l (Middle Temporal Gyrus, anterior division Left)
52 voxels (6%) covering 9% of atlas.aPaHC l (Parahippocampal Gyrus, anterior division Left)
46 voxels (5%) covering 3% of atlas.pMTG l (Middle Temporal Gyrus, posterior division Left)
31 voxels (3%) covering 4% of atlas.Hippocampus l
25 voxels (3%) covering 7% of atlas.aITG l (Inferior Temporal Gyrus, anterior division Left)
22 voxels (2%) covering 7% of atlas.aTFusC l (Temporal Fusiform Cortex, anterior division Left)
22 voxels (2%) covering 1% of atlas.Brain-Stem
17 voxels (2%) covering 5% of atlas.Amygdala l
9 voxels (1%) covering 1% of atlas.pITG l (Inferior Temporal Gyrus, posterior division Left)
6 voxels (1%) covering 2% of atlas.pPaHC l (Parahippocampal Gyrus, posterior division Left)
1 voxels (0%) covering 0% of atlas.Cereb45 l (Cerebelum 4 5 Left)
268 voxels (29%) covering 0% of atlas.not-labeled

**Table S3. SBC maps in asymptomatic carriers using 7Am as the seed.**

| Cluster (x,y,z) | Size (vox) | size p-FWE | size p-FDR | size p-unc | peak p-FWE | peak p-unc |
|---|---|---|---|---|---|---|
| -08 -62 +62 | 7257 | 0.000000 | 0.000000 | 0.000000 | 0.000001 | 0.000000 |
| +40 +34 -02 | 2247 | 0.000906 | 0.000398 | 0.000004 | 0.916014 | 0.000056 |
| -36 -20 -32 | 1014 | 0.141110 | 0.044500 | 0.000661 | 0.998966 | 0.000203 |

**Cluster -8 -62 +62**:
1825 voxels (25%) covering 33% of atlas.Precuneous (Precuneous Cortex)
1358 voxels (19%) covering 27% of atlas.sLOC l (Lateral Occipital Cortex, superior division Left)
1209 voxels (17%) covering 25% of atlas.sLOC r (Lateral Occipital Cortex, superior division Right)
589 voxels (8%) covering 40% of atlas.SPL r (Superior Parietal Lobule Right)
238 voxels (3%) covering 16% of atlas.SPL l (Superior Parietal Lobule Left)
133 voxels (2%) covering 4% of atlas.PostCG l (Postcentral Gyrus Left)
132 voxels (2%) covering 6% of atlas.PC (Cingulate Gyrus, posterior division)
98 voxels (1%) covering 3% of atlas.PostCG r (Postcentral Gyrus Right)
96 voxels (1%) covering 2% of atlas.PreCG l (Precentral Gyrus Left)
58 voxels (1%) covering 1% of atlas.PreCG r (Precentral Gyrus Right)
55 voxels (1%) covering 4% of atlas.pSMG r (Supramarginal Gyrus, posterior division Right)
21 voxels (0%) covering 3% of atlas.aSMG r (Supramarginal Gyrus, anterior division Right)
11 voxels (0%) covering 1% of atlas.iLOC r (Lateral Occipital Cortex, inferior division Right)
7 voxels (0%) covering 5% of atlas.SCC r (Supracalcarine Cortex Right)
1427 voxels (20%) covering 0% of atlas.not-labeled

**Cluster +40 +34 -2**:
175 voxels (8%) covering 32% of atlas.IFG tri r (Inferior Frontal Gyrus, pars triangularis Right)
159 voxels (7%) covering 12% of atlas.IC r (Insular Cortex Right)
115 voxels (5%) covering 14% of atlas.Putamen r
113 voxels (5%) covering 8% of atlas.FOrb r (Frontal Orbital Cortex Right)
108 voxels (5%) covering 1% of atlas.FP r (Frontal Pole Right)
100 voxels (4%) covering 4% of atlas.AC (Cingulate Gyrus, anterior division)
90 voxels (4%) covering 17% of atlas.Caudate r
85 voxels (4%) covering 6% of atlas.PaCiG r (Paracingulate Gyrus Right)
83 voxels (4%) covering 12% of atlas.IFG oper r (Inferior Frontal Gyrus, pars opercularis Right)
77 voxels (3%) covering 9% of atlas.CO r (Central Opercular Cortex Right)
41 voxels (2%) covering 13% of atlas.FO r (Frontal Operculum Cortex Right)
21 voxels (1%) covering 25% of atlas.Accumbens r
1 voxels (0%) covering 0% of atlas.SFG r (Superior Frontal Gyrus Right)
1079 voxels (48%) covering 0% of atlas.not-labeled

**Cluster -36 -20 -32**:
332 voxels (33%) covering 39% of atlas.pTFusC l (Temporal Fusiform Cortex, posterior division Left)
133 voxels (13%) covering 17% of atlas.Hippocampus l
96 voxels (9%) covering 25% of atlas.pPaHC l (Parahippocampal Gyrus, posterior division Left)
63 voxels (6%) covering 5% of atlas.Thalamus l
30 voxels (3%) covering 2% of atlas.LG l (Lingual Gyrus Left)
29 voxels (3%) covering 3% of atlas.pITG l (Inferior Temporal Gyrus, posterior division Left)
19 voxels (2%) covering 0% of atlas.Brain-Stem
5 voxels (0%) covering 1% of atlas.aPaHC l (Parahippocampal Gyrus, anterior division Left)
3 voxels (0%) covering 0% of atlas.TOFusC l (Temporal Occipital Fusiform Cortex Left)
3 voxels (0%) covering 0% of atlas.Cereb45 l (Cerebelum 4 5 Left)
301 voxels (30%) covering 0% of atlas.not-labeled

**Table S4. SBC maps in asymptomatic carriers using 7Pm as the seed.**

| Cluster (x,y,z) | Size (vox) | size p-FWE | size p-FDR | size p-unc | peak p-FWE | peak p-unc |
|---|---|---|---|---|---|---|
| -02 -68 +48 | 5444 | 0.000000 | 0.000000 | 0.000000 | 0.000025 | 0.000000 |
| -34 +00 +20 | 4401 | 0.000001 | 0.000000 | 0.000000 | 0.715511 | 0.000024 |
| +36 -62 +18 | 1056 | 0.112883 | 0.033197 | 0.000516 | 0.728755 | 0.000025 |

**Cluster -2 -68 +48**:
2483 voxels (46%) covering 44% of atlas.Precuneous (Precuneous Cortex)
773 voxels (14%) covering 16% of atlas.sLOC l (Lateral Occipital Cortex, superior division Left)
401 voxels (7%) covering 17% of atlas.PC (Cingulate Gyrus, posterior division)
209 voxels (4%) covering 4% of atlas.sLOC r (Lateral Occipital Cortex, superior division Right)
62 voxels (1%) covering 10% of atlas.Cuneal r (Cuneal Cortex Right)
49 voxels (1%) covering 2% of atlas.iLOC l (Lateral Occipital Cortex, inferior division Left)
28 voxels (1%) covering 3% of atlas.toMTG l (Middle Temporal Gyrus, temporooccipital part Left)
14 voxels (0%) covering 0% of atlas.PostCG l (Postcentral Gyrus Left)
7 voxels (0%) covering 0% of atlas.SPL r (Superior Parietal Lobule Right)
5 voxels (0%) covering 0% of atlas.SPL l (Superior Parietal Lobule Left)
3 voxels (0%) covering 2% of atlas.SCC r (Supracalcarine Cortex Right)
1 voxels (0%) covering 0% of atlas.ICC r (Intracalcarine Cortex Right)
1409 voxels (26%) covering 0% of atlas.not-labeled

**Cluster -34 +0 +20**:
441 voxels (10%) covering 33% of atlas.IC r (Insular Cortex Right)
316 voxels (7%) covering 23% of atlas.Thalamus l
252 voxels (6%) covering 20% of atlas.Thalamus r
210 voxels (5%) covering 59% of atlas.FO l (Frontal Operculum Cortex Left)
195 voxels (4%) covering 15% of atlas.IC l (Insular Cortex Left)
183 voxels (4%) covering 21% of atlas.Putamen l
160 voxels (4%) covering 51% of atlas.FO r (Frontal Operculum Cortex Right)
119 voxels (3%) covering 14% of atlas.CO r (Central Opercular Cortex Right)
104 voxels (2%) covering 7% of atlas.FOrb r (Frontal Orbital Cortex Right)
103 voxels (2%) covering 13% of atlas.Putamen r
100 voxels (2%) covering 33% of atlas.Pallidum l
97 voxels (2%) covering 36% of atlas.Pallidum r
95 voxels (2%) covering 15% of atlas.IFG tri l (Inferior Frontal Gyrus, pars triangularis Left)
91 voxels (2%) covering 9% of atlas.CO l (Central Opercular Cortex Left)
77 voxels (2%) covering 1% of atlas.FP l (Frontal Pole Left)
70 voxels (2%) covering 83% of atlas.Accumbens r
64 voxels (1%) covering 17% of atlas.PP r (Planum Polare Right)
63 voxels (1%) covering 9% of atlas.IFG oper r (Inferior Frontal Gyrus, pars opercularis Right)
59 voxels (1%) covering 11% of atlas.IFG tri r (Inferior Frontal Gyrus, pars triangularis Right)
58 voxels (1%) covering 11% of atlas.Caudate l
37 voxels (1%) covering 5% of atlas.Hippocampus r
27 voxels (1%) covering 5% of atlas.Caudate r
12 voxels (0%) covering 11% of atlas.Accumbens l
8 voxels (0%) covering 0% of atlas.PreCG r (Precentral Gyrus Right)
6 voxels (0%) covering 0% of atlas.TP r (Temporal Pole Right)
5 voxels (0%) covering 1% of atlas.IFG oper l (Inferior Frontal Gyrus, pars opercularis Left)
4 voxels (0%) covering 0% of atlas.FP r (Frontal Pole Right)
4 voxels (0%) covering 0% of atlas.FOrb l (Frontal Orbital Cortex Left)
4 voxels (0%) covering 1% of atlas.Amygdala r
1 voxels (0%) covering 0% of atlas.aSTG r (Superior Temporal Gyrus, anterior division Right)

1436 voxels (33%) covering 0% of atlas.not-labeled

**Cluster +36 -62 +18**:
711 voxels (67%) covering 15% of atlas.sLOC r (Lateral Occipital Cortex, superior division Right)
110 voxels (10%) covering 5% of atlas.iLOC r (Lateral Occipital Cortex, inferior division Right)
64 voxels (6%) covering 6% of atlas.toMTG r (Middle Temporal Gyrus, temporooccipital part Right)
15 voxels (1%) covering 1% of atlas.AG r (Angular Gyrus Right)
2 voxels (0%) covering 0% of atlas.Precuneous (Precuneous Cortex)
154 voxels (15%) covering 0% of atlas.not-labeled

**Table S5. SBC maps in asymptomatic carriers using 7m as the seed.**

| Cluster (x,y,z) | size | size p-FWE | size p-FDR | size p-unc | peak p-FWE | peak p-unc |
|---|---|---|---|---|---|---|
| +52 -60 +14 | 5723 | 0.000000 | 0.000000 | 0.000000 | 0.082592 | 0.000001 |
| +02 -60 +30 | 5431 | 0.000000 | 0.000000 | 0.000000 | 0.000019 | 0.000000 |
| -44 -62 +16 | 4091 | 0.000002 | 0.000000 | 0.000000 | 0.772239 | 0.000029 |
| -36 +32 +20 | 2433 | 0.000382 | 0.000078 | 0.000002 | 1.000000 | 0.000705 |
| -04 +20 -20 | 1243 | 0.045852 | 0.007667 | 0.000200 | 0.999821 | 0.000265 |
| -14 +30 +54 | 1122 | 0.079054 | 0.011210 | 0.000350 | 1.000000 | 0.000838 |
| -54 -36 +36 | 870 | 0.247912 | 0.033240 | 0.001212 | 0.951785 | 0.000070 |

**Cluster +52 -60 +14**:
804 voxels (14%) covering 59% of atlas.pMTG r (Middle Temporal Gyrus, posterior division Right)
788 voxels (14%) covering 33% of atlas.TP r (Temporal Pole Right)
559 voxels (10%) covering 48% of atlas.toMTG r (Middle Temporal Gyrus, temporooccipital part Right)
408 voxels (7%) covering 28% of atlas.AG r (Angular Gyrus Right)
316 voxels (6%) covering 77% of atlas.aMTG r (Middle Temporal Gyrus, anterior division Right)
316 voxels (6%) covering 7% of atlas.sLOC r (Lateral Occipital Cortex, superior division Right)
200 voxels (3%) covering 10% of atlas.iLOC r (Lateral Occipital Cortex, inferior division Right)
194 voxels (3%) covering 47% of atlas.pSTG r (Superior Temporal Gyrus, posterior division Right)
164 voxels (3%) covering 13% of atlas.pSMG r (Supramarginal Gyrus, posterior division Right)
85 voxels (1%) covering 12% of atlas.pTFusC r (Temporal Fusiform Cortex, posterior division Right)
63 voxels (1%) covering 23% of atlas.aSTG r (Superior Temporal Gyrus, anterior division Right)
58 voxels (1%) covering 6% of atlas.pITG r (Inferior Temporal Gyrus, posterior division Right)
52 voxels (1%) covering 16% of atlas.aITG r (Inferior Temporal Gyrus, anterior division Right)
46 voxels (1%) covering 16% of atlas.aTFusC r (Temporal Fusiform Cortex, anterior division Right)
36 voxels (1%) covering 0% of atlas.FP r (Frontal Pole Right)
29 voxels (1%) covering 2% of atlas.FOrb r (Frontal Orbital Cortex Right)
11 voxels (0%) covering 2% of atlas.aPaHC r (Parahippocampal Gyrus, anterior division Right)
3 voxels (0%) covering 1% of atlas.PP r (Planum Polare Right)
1591 voxels (28%) covering 0% of atlas.not-labeled

**Cluster +2 -60 +30**:
2640 voxels (49%) covering 47% of atlas.Precuneous (Precuneous Cortex)
797 voxels (15%) covering 33% of atlas.PC (Cingulate Gyrus, posterior division)
244 voxels (4%) covering 38% of atlas.Cuneal r (Cuneal Cortex Right)
179 voxels (3%) covering 34% of atlas.Cuneal l (Cuneal Cortex Left)
44 voxels (1%) covering 31% of atlas.SCC r (Supracalcarine Cortex Right)
17 voxels (0%) covering 3% of atlas.Ver45 (Vermis 4 5)

16 voxels (0%) covering 22% of atlas.SCC l (Supracalcarine Cortex Left)
15 voxels (0%) covering 2% of atlas.ICC r (Intracalcarine Cortex Right)
11 voxels (0%) covering 2% of atlas.ICC l (Intracalcarine Cortex Left)
5 voxels (0%) covering 0% of atlas.sLOC r (Lateral Occipital Cortex, superior division Right)
2 voxels (0%) covering 0% of atlas.LG l (Lingual Gyrus Left)
2 voxels (0%) covering 0% of atlas.Thalamus l
1 voxels (0%) covering 0% of atlas.sLOC l (Lateral Occipital Cortex, superior division Left)
1458 voxels (27%) covering 0% of atlas.not-labeled

**Cluster -44 -62 +16**:
645 voxels (16%) covering 13% of atlas.sLOC l (Lateral Occipital Cortex, superior division Left)
572 voxels (14%) covering 41% of atlas.pMTG l (Middle Temporal Gyrus, posterior division Left)
294 voxels (7%) covering 12% of atlas.TP l (Temporal Pole Left)
290 voxels (7%) covering 31% of atlas.AG l (Angular Gyrus Left)
198 voxels (5%) covering 10% of atlas.iLOC l (Lateral Occipital Cortex, inferior division Left)
197 voxels (5%) covering 23% of atlas.toMTG l (Middle Temporal Gyrus, temporooccipital part Left)
140 voxels (3%) covering 36% of atlas.pSTG l (Superior Temporal Gyrus, posterior division Left)
135 voxels (3%) covering 16% of atlas.pTFusC l (Temporal Fusiform Cortex, posterior division Left)
131 voxels (3%) covering 29% of atlas.aMTG l (Middle Temporal Gyrus, anterior division Left)
91 voxels (2%) covering 9% of atlas.pSMG l (Supramarginal Gyrus, posterior division Left)
91 voxels (2%) covering 16% of atlas.aPaHC l (Parahippocampal Gyrus, anterior division Left)
85 voxels (2%) covering 25% of atlas.aITG l (Inferior Temporal Gyrus, anterior division Left)
75 voxels (2%) covering 10% of atlas.Hippocampus l
54 voxels (1%) covering 17% of atlas.aTFusC l (Temporal Fusiform Cortex, anterior division Left)
51 voxels (1%) covering 5% of atlas.pITG l (Inferior Temporal Gyrus, posterior division Left)
19 voxels (0%) covering 3% of atlas.PT l (Planum Temporale Left)
15 voxels (0%) covering 5% of atlas.Amygdala l
10 voxels (0%) covering 3% of atlas.pPaHC l (Parahippocampal Gyrus, posterior division Left)
9 voxels (0%) covering 3% of atlas.aSTG l (Superior Temporal Gyrus, anterior division Left)
7 voxels (0%) covering 2% of atlas.HG l (Heschl's Gyrus Left)
982 voxels (24%) covering 0% of atlas.not-labeled

**Cluster -36 +32 +20**:
302 voxels (12%) covering 10% of atlas.MidFG l (Middle Frontal Gyrus Left)
191 voxels (8%) covering 14% of atlas.IC l (Insular Cortex Left)
170 voxels (7%) covering 13% of atlas.Thalamus r
115 voxels (5%) covering 3% of atlas.PreCG l (Precentral Gyrus Left)
113 voxels (5%) covering 8% of atlas.Thalamus l
98 voxels (4%) covering 4% of atlas.AC (Cingulate Gyrus, anterior division)
47 voxels (2%) covering 9% of atlas.Caudate l
39 voxels (2%) covering 4% of atlas.CO l (Central Opercular Cortex Left)
33 voxels (1%) covering 0% of atlas.FP l (Frontal Pole Left)
31 voxels (1%) covering 2% of atlas.PaCiG l (Paracingulate Gyrus Left)
23 voxels (1%) covering 7% of atlas.Amygdala l
21 voxels (1%) covering 1% of atlas.TP l (Temporal Pole Left)
16 voxels (1%) covering 2% of atlas.Putamen l
11 voxels (0%) covering 3% of atlas.FO l (Frontal Operculum Cortex Left)
5 voxels (0%) covering 1% of atlas.Putamen r
3 voxels (0%) covering 1% of atlas.PP l (Planum Polare Left)
3 voxels (0%) covering 1% of atlas.Caudate r

2 voxels (0%) covering 0% of atlas.IFG tri l (Inferior Frontal Gyrus, pars triangularis Left)
1210 voxels (50%) covering 0% of atlas.not-labeled

**Cluster -4 +20 -20**:
387 voxels (31%) covering 5% of atlas.FP r (Frontal Pole Right)
291 voxels (23%) covering 30% of atlas.MedFC (Frontal Medial Cortex)
229 voxels (18%) covering 3% of atlas.FP l (Frontal Pole Left)
80 voxels (6%) covering 7% of atlas.SubCalC (Subcallosal Cortex)
4 voxels (0%) covering 0% of atlas.PaCiG l (Paracingulate Gyrus Left)
1 voxels (0%) covering 0% of atlas.FOrb l (Frontal Orbital Cortex Left)
251 voxels (20%) covering 0% of atlas.not-labeled

**Cluster -14 +30 +54**:
395 voxels (35%) covering 14% of atlas.SFG l (Superior Frontal Gyrus Left)
177 voxels (16%) covering 7% of atlas.SFG r (Superior Frontal Gyrus Right)
113 voxels (10%) covering 2% of atlas.FP l (Frontal Pole Left)
103 voxels (9%) covering 1% of atlas.FP r (Frontal Pole Right)
21 voxels (2%) covering 2% of atlas.PaCiG r (Paracingulate Gyrus Right)
11 voxels (1%) covering 1% of atlas.PaCiG l (Paracingulate Gyrus Left)
8 voxels (1%) covering 0% of atlas.MidFG l (Middle Frontal Gyrus Left)
294 voxels (26%) covering 0% of atlas.not-labeled

**Cluster -54 -36 +36**:
448 voxels (51%) covering 47% of atlas.aSMG l (Supramarginal Gyrus, anterior division Left)
104 voxels (12%) covering 18% of atlas.PO l (Parietal Operculum Cortex Left)
83 voxels (10%) covering 8% of atlas.CO l (Central Opercular Cortex Left)
40 voxels (5%) covering 1% of atlas.PostCG l (Postcentral Gyrus Left)
6 voxels (1%) covering 1% of atlas.pSMG l (Supramarginal Gyrus, posterior division Left)
189 voxels (22%) covering 0% of atlas.not-labeled

**Table S6. SBC maps in asymptomatic carriers using PCV as the seed.**

| Cluster (x,y,z) | Size (vox) | size p-FWE | size p-FDR | size p-unc | peak p-FWE | peak p-unc |
|---|---|---|---|---|---|---|
| -04 -46 +48 | 8334 | 0.000000 | 0.000000 | 0.000000 | 0.008748 | 0.000000 |
| +32 +26 +12 | 2949 | 0.000116 | 0.000049 | 0.000001 | 0.876524 | 0.000048 |

**Cluster -4 -46 +48**:
2564 voxels (31%) covering 46% of atlas.Precuneous (Precuneous Cortex)
377 voxels (5%) covering 16% of atlas.PC (Cingulate Gyrus, posterior division)
371 voxels (4%) covering 7% of atlas.sLOC l (Lateral Occipital Cortex, superior division Left)
332 voxels (4%) covering 9% of atlas.PostCG l (Postcentral Gyrus Left)
320 voxels (4%) covering 37% of atlas.pTFusC l (Temporal Fusiform Cortex, posterior division Left)
251 voxels (3%) covering 8% of atlas.PostCG r (Postcentral Gyrus Right)
251 voxels (3%) covering 5% of atlas.sLOC r (Lateral Occipital Cortex, superior division Right)
190 voxels (2%) covering 25% of atlas.Hippocampus l
119 voxels (1%) covering 31% of atlas.pPaHC l (Parahippocampal Gyrus, posterior division Left)
102 voxels (1%) covering 7% of atlas.SPL r (Superior Parietal Lobule Right)
95 voxels (1%) covering 5% of atlas.iLOC r (Lateral Occipital Cortex, inferior division Right)
92 voxels (1%) covering 14% of atlas.Cuneal r (Cuneal Cortex Right)

80 voxels (1%) covering 14% of atlas.aPaHC l (Parahippocampal Gyrus, anterior division Left)
61 voxels (1%) covering 1% of atlas.PreCG l (Precentral Gyrus Left)
59 voxels (1%) covering 5% of atlas.toMTG r (Middle Temporal Gyrus, temporooccipital part Right)
54 voxels (1%) covering 1% of atlas.Brain-Stem
43 voxels (1%) covering 3% of atlas.SPL l (Superior Parietal Lobule Left)
42 voxels (1%) covering 5% of atlas.Cereb45 l (Cerebelum 4 5 Left)
39 voxels (0%) covering 8% of atlas.Cuneal l (Cuneal Cortex Left)
31 voxels (0%) covering 2% of atlas.Thalamus l
26 voxels (0%) covering 8% of atlas.Amygdala l
22 voxels (0%) covering 1% of atlas.PreCG r (Precentral Gyrus Right)
17 voxels (0%) covering 12% of atlas.SCC r (Supracalcarine Cortex Right)
16 voxels (0%) covering 1% of atlas.AG r (Angular Gyrus Right)
15 voxels (0%) covering 1% of atlas.iLOC l (Lateral Occipital Cortex, inferior division Left)
15 voxels (0%) covering 5% of atlas.aTFusC l (Temporal Fusiform Cortex, anterior division Left)
12 voxels (0%) covering 1% of atlas.LG l (Lingual Gyrus Left)
9 voxels (0%) covering 1% of atlas.ICC l (Intracalcarine Cortex Left)
9 voxels (0%) covering 12% of atlas.SCC l (Supracalcarine Cortex Left)
5 voxels (0%) covering 0% of atlas.pITG l (Inferior Temporal Gyrus, posterior division Left)
3 voxels (0%) covering 0% of atlas.TP l (Temporal Pole Left)
3 voxels (0%) covering 0% of atlas.ICC r (Intracalcarine Cortex Right)
1 voxels (0%) covering 0% of atlas.LG r (Lingual Gyrus Right)
1 voxels (0%) covering 0% of atlas.TOFusC l (Temporal Occipital Fusiform Cortex Left)
2707 voxels (32%) covering 1% of atlas.not-labeled

**Cluster +32 +26 +12**:
524 voxels (18%) covering 6% of atlas.FP r (Frontal Pole Right)
371 voxels (13%) covering 28% of atlas.IC r (Insular Cortex Right)
250 voxels (8%) covering 36% of atlas.IFG oper r (Inferior Frontal Gyrus, pars opercularis Right)
241 voxels (8%) covering 77% of atlas.FO r (Frontal Operculum Cortex Right)
145 voxels (5%) covering 3% of atlas.PreCG r (Precentral Gyrus Right)
129 voxels (4%) covering 23% of atlas.IFG tri r (Inferior Frontal Gyrus, pars triangularis Right)
104 voxels (4%) covering 12% of atlas.CO r (Central Opercular Cortex Right)
44 voxels (1%) covering 2% of atlas.AC (Cingulate Gyrus, anterior division)
31 voxels (1%) covering 2% of atlas.FOrb r (Frontal Orbital Cortex Right)
27 voxels (1%) covering 1% of atlas.TP r (Temporal Pole Right)
25 voxels (1%) covering 3% of atlas.Putamen r
12 voxels (0%) covering 0% of atlas.MidFG r (Middle Frontal Gyrus Right)
3 voxels (0%) covering 1% of atlas.Amygdala r
2 voxels (0%) covering 0% of atlas.PaCiG r (Paracingulate Gyrus Right)
2 voxels (0%) covering 0% of atlas.Hippocampus r
1039 voxels (35%) covering 0% of atlas.not-labeled

**Table S7. SBC maps in asymptomatic carriers using POS2 as the seed.**

| Cluster (x,y,z) | Size (vox) | size p-FWE | size p-FDR | size p-unc | peak p-FWE | peak p-unc |
|---|---|---|---|---|---|---|
| +08 -68 +36 | 5422 | 0.000000 | 0.000000 | 0.000000 | 0.002795 | 0.000000 |

**Cluster +8 -68 +36**:
1526 voxels (28%) covering 27% of atlas.Precuneous (Precuneous Cortex)
694 voxels (13%) covering 29% of atlas.PC (Cingulate Gyrus, posterior division)

450 voxels (8%) covering 70% of atlas.Cuneal r (Cuneal Cortex Right)
339 voxels (6%) covering 65% of atlas.Cuneal l (Cuneal Cortex Left)
313 voxels (6%) covering 7% of atlas.sLOC r (Lateral Occipital Cortex, superior division Right)
64 voxels (1%) covering 1% of atlas.sLOC l (Lateral Occipital Cortex, superior division Left)
44 voxels (1%) covering 3% of atlas.AG r (Angular Gyrus Right)
30 voxels (1%) covering 21% of atlas.SCC r (Supracalcarine Cortex Right)
24 voxels (0%) covering 2% of atlas.SPL r (Superior Parietal Lobule Right)
21 voxels (0%) covering 1% of atlas.AC (Cingulate Gyrus, anterior division)
8 voxels (0%) covering 1% of atlas.pSMG r (Supramarginal Gyrus, posterior division Right)
3 voxels (0%) covering 0% of atlas.ICC r (Intracalcarine Cortex Right)
3 voxels (0%) covering 0% of atlas.Ver45 (Vermis 4 5)
2 voxels (0%) covering 0% of atlas.Thalamus l
1 voxels (0%) covering 0% of atlas.ICC l (Intracalcarine Cortex Left)
1900 voxels (35%) covering 1% of atlas.not-labeled

**Table S8. SBC maps differences between aC and nC using the whole Precuneus as the seed.**

| Cluster (x,y,z) | Size (vox) | size p-FWE | size p-FDR | size p-unc | peak p-FWE | peak p-unc |
|---|---|---|---|---|---|---|
| +46 -70 -20 | 2765 | 0.000114 | 0.000111 | 0.000000 | 0.480123 | 0.000011 |
| +20 +48 +08 | 2208 | 0.000861 | 0.000420 | 0.000004 | 0.891091 | 0.000047 |
| -16 -76 -20 | 1049 | 0.108950 | 0.037517 | 0.000489 | 0.999983 | 0.000360 |

**Cluster +46 -70 -20**:
731 voxels (26%) covering 36% of atlas.iLOC r (Lateral Occipital Cortex, inferior division Right)
360 voxels (13%) covering 14% of atlas.OP r (Occipital Pole Right)
292 voxels (11%) covering 12% of atlas.Cereb1 r (Cerebelum Crus1 Right)
254 voxels (9%) covering 29% of atlas.OFusG r (Occipital Fusiform Gyrus Right)
241 voxels (9%) covering 30% of atlas.TOFusC r (Temporal Occipital Fusiform Cortex Right)
194 voxels (7%) covering 25% of atlas.toITG r (Inferior Temporal Gyrus, temporooccipital part Right)
129 voxels (5%) covering 7% of atlas.LG r (Lingual Gyrus Right)
83 voxels (3%) covering 7% of atlas.toMTG r (Middle Temporal Gyrus, temporooccipital part Right)
56 voxels (2%) covering 4% of atlas.Cereb6 r (Cerebelum 6 Right)
1 voxels (0%) covering 0% of atlas.pTFusC r (Temporal Fusiform Cortex, posterior division Right)
424 voxels (15%) covering 0% of atlas.not-labeled

**Cluster +20 +48 +8**:
465 voxels (21%) covering 18% of atlas.AC (Cingulate Gyrus, anterior division)
282 voxels (13%) covering 29% of atlas.MedFC (Frontal Medial Cortex)
195 voxels (9%) covering 2% of atlas.FP r (Frontal Pole Right)
105 voxels (5%) covering 8% of atlas.PaCiG l (Paracingulate Gyrus Left)
104 voxels (5%) covering 2% of atlas.FP l (Frontal Pole Left)
74 voxels (3%) covering 5% of atlas.PaCiG r (Paracingulate Gyrus Right)
25 voxels (1%) covering 2% of atlas.SubCalC (Subcallosal Cortex)
958 voxels (43%) covering 0% of atlas.not-labeled

**Cluster -16 -76 -20**:
314 voxels (30%) covering 15% of atlas.iLOC l (Lateral Occipital Cortex, inferior division Left)
292 voxels (28%) covering 19% of atlas.LG l (Lingual Gyrus Left)

105 voxels (10%) covering 11% of atlas.OFusG l (Occipital Fusiform Gyrus Left)
46 voxels (4%) covering 4% of atlas.Cereb6 l (Cerebelum 6 Left)
15 voxels (1%) covering 1% of atlas.Cereb1 l (Cerebelum Crus1 Left)
6 voxels (1%) covering 1% of atlas.Ver45 (Vermis 4 5)
2 voxels (0%) covering 0% of atlas.Cereb45 l (Cerebelum 4 5 Left)
1 voxels (0%) covering 0% of atlas.toITG l (Inferior Temporal Gyrus, temporooccipital part Left)
1 voxels (0%) covering 0% of atlas.sLOC l (Lateral Occipital Cortex, superior division Left)
1 voxels (0%) covering 0% of atlas.ICC l (Intracalcarine Cortex Left)
1 voxels (0%) covering 0% of atlas.OP l (Occipital Pole Left)
265 voxels (25%) covering 0% of atlas.not-labeled

**Table S9. SBC maps differences between aC and nC using 7m as the seed.**

| Cluster (x,y,z) | size | size p-FWE | size p-FDR | size p-unc | peak p-FWE | peak p-unc |
|---|---|---|---|---|---|---|
| -14 +40 -04 | 1516 | 0.013959 | 0.013335 | 0.000060 | 0.782710 | 0.000030 |
| +16 -92 -02 | 1285 | 0.038040 | 0.018394 | 0.000165 | 1.000000 | 0.001763 |

**Cluster -14 +40 -4**:
413 voxels (27%) covering 42% of atlas.MedFC (Frontal Medial Cortex)
171 voxels (11%) covering 7% of atlas.AC (Cingulate Gyrus, anterior division)
128 voxels (8%) covering 10% of atlas.PaCiG l (Paracingulate Gyrus Left)
81 voxels (5%) covering 1% of atlas.FP l (Frontal Pole Left)
57 voxels (4%) covering 1% of atlas.FP r (Frontal Pole Right)
40 voxels (3%) covering 3% of atlas.PaCiG r (Paracingulate Gyrus Right)
19 voxels (1%) covering 2% of atlas.SubCalC (Subcallosal Cortex)
1 voxels (0%) covering 0% of atlas.FOrb r (Frontal Orbital Cortex Right)
606 voxels (40%) covering 0% of atlas.not-labeled

**Cluster +16 -92 -2**:
388 voxels (30%) covering 16% of atlas.OP r (Occipital Pole Right)
230 voxels (18%) covering 9% of atlas.Cereb1 r (Cerebelum Crus1 Right)
118 voxels (9%) covering 7% of atlas.LG r (Lingual Gyrus Right)
118 voxels (9%) covering 13% of atlas.OFusG r (Occipital Fusiform Gyrus Right)
103 voxels (8%) covering 5% of atlas.iLOC r (Lateral Occipital Cortex, inferior division Right)
2 voxels (0%) covering 0% of atlas.Cereb2 l (Cerebelum Crus2 Left)
2 voxels (0%) covering 1% of atlas.Ver6 (Vermis 6)
1 voxels (0%) covering 0% of atlas.sLOC r (Lateral Occipital Cortex, superior division Right)
323 voxels (25%) covering 0% of atlas.not-labeled

**Table S10. SBC maps differences between aC and nC using 7Am as the seed.**

| Cluster (x,y,z) | size | size p-FWE | size p-FDR | size p-unc | peak p-FWE | peak p-unc |
|---|---|---|---|---|---|---|
| -38 -66 +00 | 2413 | 0.000494 | 0.000249 | 0.000002 | 0.999928 | 0.000311 |
| -08 -60 +24 | 2378 | 0.000561 | 0.000249 | 0.000002 | 0.990597 | 0.000124 |
| +48 -46 +10 | 2105 | 0.001538 | 0.000455 | 0.000007 | 0.999765 | 0.000261 |
| -20 +22 +44 | 1260 | 0.047444 | 0.010770 | 0.000211 | 0.621579 | 0.000018 |
| +34 -12 +08 | 1022 | 0.136166 | 0.025480 | 0.000636 | 0.998676 | 0.000194 |
| +36 -20 +42 | 988 | 0.158432 | 0.025480 | 0.000749 | 1.000000 | 0.001439 |
| -36 -78 +38 | 929 | 0.205772 | 0.029170 | 0.001001 | 1.000000 | 0.000767 |

| | | | | | | |
|---|---|---|---|---|---|---|
| -20 +60 +02 | 780 | 0.388923 | 0.049017 | 0.002140 | 1.000000 | 0.000625 |
| -12 -70 -12 | 778 | 0.392095 | 0.049017 | 0.002163 | 1.000000 | 0.000691 |

**Cluster -38 -66 +0**:
194 voxels (8%) covering 20% of atlas.CO l (Central Opercular Cortex Left)
163 voxels (7%) covering 7% of atlas.Cereb1 l (Cerebelum Crus1 Left)
159 voxels (7%) covering 24% of atlas.TOFusC l (Temporal Occipital Fusiform Cortex Left)
144 voxels (6%) covering 17% of atlas.toMTG l (Middle Temporal Gyrus, temporooccipital part Left)
128 voxels (5%) covering 12% of atlas.pSMG l (Supramarginal Gyrus, posterior division Left)
110 voxels (5%) covering 5% of atlas.iLOC l (Lateral Occipital Cortex, inferior division Left)
90 voxels (4%) covering 12% of atlas.IFG oper l (Inferior Frontal Gyrus, pars opercularis Left)
88 voxels (4%) covering 9% of atlas.OFusG l (Occipital Fusiform Gyrus Left)
83 voxels (3%) covering 23% of atlas.FO l (Frontal Operculum Cortex Left)
82 voxels (3%) covering 2% of atlas.PreCG l (Precentral Gyrus Left)
80 voxels (3%) covering 29% of atlas.aSTG l (Superior Temporal Gyrus, anterior division Left)
63 voxels (3%) covering 5% of atlas.pMTG l (Middle Temporal Gyrus, posterior division Left)
57 voxels (2%) covering 10% of atlas.PT l (Planum Temporale Left)
53 voxels (2%) covering 17% of atlas.HG l (Heschl's Gyrus Left)
49 voxels (2%) covering 5% of atlas.AG l (Angular Gyrus Left)
48 voxels (2%) covering 4% of atlas.IC l (Insular Cortex Left)
46 voxels (2%) covering 13% of atlas.PP l (Planum Polare Left)
35 voxels (1%) covering 6% of atlas.PO l (Parietal Operculum Cortex Left)
34 voxels (1%) covering 1% of atlas.TP l (Temporal Pole Left)
26 voxels (1%) covering 7% of atlas.pSTG l (Superior Temporal Gyrus, posterior division Left)
24 voxels (1%) covering 3% of atlas.pTFusC l (Temporal Fusiform Cortex, posterior division Left)
16 voxels (1%) covering 2% of atlas.aSMG l (Supramarginal Gyrus, anterior division Left)
14 voxels (1%) covering 1% of atlas.Cereb6 l (Cerebelum 6 Left)
4 voxels (0%) covering 1% of atlas.toITG l (Inferior Temporal Gyrus, temporooccipital part Left)
3 voxels (0%) covering 0% of atlas.sLOC l (Lateral Occipital Cortex, superior division Left)
2 voxels (0%) covering 0% of atlas.PostCG l (Postcentral Gyrus Left)
618 voxels (26%) covering 0% of atlas.not-labeled

**Cluster -8 -60 +24**:
1327 voxels (56%) covering 24% of atlas.Precuneous (Precuneous Cortex)
704 voxels (30%) covering 29% of atlas.PC (Cingulate Gyrus, posterior division)
30 voxels (1%) covering 5% of atlas.Ver45 (Vermis 4 5)
5 voxels (0%) covering 1% of atlas.Cereb45 l (Cerebelum 4 5 Left)
1 voxels (0%) covering 0% of atlas.Cuneal l (Cuneal Cortex Left)
311 voxels (13%) covering 0% of atlas.not-labeled

**Cluster +48 -46 +10**:
317 voxels (15%) covering 16% of atlas.iLOC r (Lateral Occipital Cortex, inferior division Right)
233 voxels (11%) covering 20% of atlas.toMTG r (Middle Temporal Gyrus, temporooccipital part Right)
170 voxels (8%) covering 4% of atlas.PreCG r (Precentral Gyrus Right)
108 voxels (5%) covering 12% of atlas.CO r (Central Opercular Cortex Right)
107 voxels (5%) covering 16% of atlas.IFG oper r (Inferior Frontal Gyrus, pars opercularis Right)
92 voxels (4%) covering 1% of atlas.FP r (Frontal Pole Right)
86 voxels (4%) covering 4% of atlas.TP r (Temporal Pole Right)
80 voxels (4%) covering 6% of atlas.pMTG r (Middle Temporal Gyrus, posterior division Right)

46 voxels (2%) covering 17% of atlas.aSTG r (Superior Temporal Gyrus, anterior division Right)
43 voxels (2%) covering 11% of atlas.PP r (Planum Polare Right)
43 voxels (2%) covering 15% of atlas.HG r (Heschl's Gyrus Right)
41 voxels (2%) covering 3% of atlas.AG r (Angular Gyrus Right)
36 voxels (2%) covering 1% of atlas.Cereb1 r (Cerebelum Crus1 Right)
35 voxels (2%) covering 8% of atlas.pSTG r (Superior Temporal Gyrus, posterior division Right)
18 voxels (1%) covering 2% of atlas.OFusG r (Occipital Fusiform Gyrus Right)
16 voxels (1%) covering 1% of atlas.pSMG r (Supramarginal Gyrus, posterior division Right)
13 voxels (1%) covering 2% of atlas.PO r (Parietal Operculum Cortex Right)
12 voxels (1%) covering 3% of atlas.PT r (Planum Temporale Right)
9 voxels (0%) covering 2% of atlas.IFG tri r (Inferior Frontal Gyrus, pars triangularis Right)
3 voxels (0%) covering 0% of atlas.OP r (Occipital Pole Right)
597 voxels (28%) covering 0% of atlas.not-labeled

**Cluster -20 +22 +44**:
392 voxels (31%) covering 13% of atlas.MidFG l (Middle Frontal Gyrus Left)
385 voxels (31%) covering 14% of atlas.SFG l (Superior Frontal Gyrus Left)
33 voxels (3%) covering 3% of atlas.PaCiG l (Paracingulate Gyrus Left)
450 voxels (36%) covering 0% of atlas.not-labeled

**Cluster +34 -12 +8**:
310 voxels (30%) covering 23% of atlas.IC r (Insular Cortex Right)
155 voxels (15%) covering 7% of atlas.TP r (Temporal Pole Right)
104 voxels (10%) covering 32% of atlas.aITG r (Inferior Temporal Gyrus, anterior division Right)
53 voxels (5%) covering 14% of atlas.PP r (Planum Polare Right)
25 voxels (2%) covering 9% of atlas.aSTG r (Superior Temporal Gyrus, anterior division Right)
19 voxels (2%) covering 2% of atlas.Putamen r
11 voxels (1%) covering 3% of atlas.Amygdala r
10 voxels (1%) covering 1% of atlas.pITG r (Inferior Temporal Gyrus, posterior division Right)
7 voxels (1%) covering 0% of atlas.FOrb r (Frontal Orbital Cortex Right)
7 voxels (1%) covering 2% of atlas.aTFusC r (Temporal Fusiform Cortex, anterior division Right)
6 voxels (1%) covering 1% of atlas.aPaHC r (Parahippocampal Gyrus, anterior division Right)
2 voxels (0%) covering 0% of atlas.aMTG r (Middle Temporal Gyrus, anterior division Right)
2 voxels (0%) covering 0% of atlas.CO r (Central Opercular Cortex Right)
311 voxels (30%) covering 0% of atlas.not-labeled

**Cluster +36 -20 +42**:
326 voxels (33%) covering 8% of atlas.PreCG r (Precentral Gyrus Right)
266 voxels (27%) covering 8% of atlas.PostCG r (Postcentral Gyrus Right)
86 voxels (9%) covering 11% of atlas.aSMG r (Supramarginal Gyrus, anterior division Right)
78 voxels (8%) covering 5% of atlas.SPL r (Superior Parietal Lobule Right)
45 voxels (5%) covering 4% of atlas.pSMG r (Supramarginal Gyrus, posterior division Right)
19 voxels (2%) covering 1% of atlas.MidFG r (Middle Frontal Gyrus Right)
168 voxels (17%) covering 0% of atlas.not-labeled

**Cluster -36 -78 +38**:
807 voxels (87%) covering 16% of atlas.sLOC l (Lateral Occipital Cortex, superior division Left)
45 voxels (5%) covering 5% of atlas.AG l (Angular Gyrus Left)
77 voxels (8%) covering 0% of atlas.not-labeled

**Cluster -20 +60 +2**:

284 voxels (36%) covering 4% of atlas.FP l (Frontal Pole Left)
142 voxels (18%) covering 2% of atlas.FP r (Frontal Pole Right)
71 voxels (9%) covering 5% of atlas.PaCiG l (Paracingulate Gyrus Left)
47 voxels (6%) covering 3% of atlas.PaCiG r (Paracingulate Gyrus Right)
41 voxels (5%) covering 4% of atlas.MedFC (Frontal Medial Cortex)
2 voxels (0%) covering 0% of atlas.AC (Cingulate Gyrus, anterior division)
1 voxels (0%) covering 0% of atlas.SFG r (Superior Frontal Gyrus Right)
192 voxels (25%) covering 0% of atlas.not-labeled

**Cluster -12 -70 -12**:
361 voxels (46%) covering 24% of atlas.LG l (Lingual Gyrus Left)
126 voxels (16%) covering 10% of atlas.Cereb6 l (Cerebelum 6 Left)
114 voxels (15%) covering 12% of atlas.OFusG l (Occipital Fusiform Gyrus Left)
21 voxels (3%) covering 1% of atlas.Cereb1 l (Cerebelum Crus1 Left)
12 voxels (2%) covering 1% of atlas.iLOC l (Lateral Occipital Cortex, inferior division Left)
8 voxels (1%) covering 1% of atlas.Cereb45 l (Cerebelum 4 5 Left)
136 voxels (17%) covering 0% of atlas.not-labeled

**Table S11. SBC maps differences between aC and nC using 7Pm as the seed.**

| Cluster (x,y,z) | size | size p-FWE | size p-FDR | size p-unc | peak p-FWE | peak p-unc |
|---|---|---|---|---|---|---|
| -18 -76 +00 | 1254 | 0.046666 | 0.049411 | 0.000206 | 1.000000 | 0.001696 |

**Cluster -18 -76 +0**:
376 voxels (30%) covering 14% of atlas.OP l (Occipital Pole Left)
343 voxels (27%) covering 23% of atlas.LG l (Lingual Gyrus Left)
85 voxels (7%) covering 3% of atlas.OP r (Occipital Pole Right)
83 voxels (7%) covering 13% of atlas.ICC l (Intracalcarine Cortex Left)
59 voxels (5%) covering 6% of atlas.OFusG l (Occipital Fusiform Gyrus Left)
40 voxels (3%) covering 8% of atlas.Cuneal l (Cuneal Cortex Left)
35 voxels (3%) covering 3% of atlas.Cereb6 l (Cerebelum 6 Left)
13 voxels (1%) covering 9% of atlas.SCC r (Supracalcarine Cortex Right)
10 voxels (1%) covering 14% of atlas.SCC l (Supracalcarine Cortex Left)
2 voxels (0%) covering 0% of atlas.Cuneal r (Cuneal Cortex Right)
1 voxels (0%) covering 0% of atlas.ICC r (Intracalcarine Cortex Right)
1 voxels (0%) covering 0% of atlas.Precuneous (Precuneous Cortex)
206 voxels (16%) covering 0% of atlas.not-labeled

**Table S12. SBC maps differences between aC and nC using PCV as the seed.**

| Cluster (x,y,z) | size | size p-FWE | size p-FDR | size p-unc | peak p-FWE | peak p-unc |
|---|---|---|---|---|---|---|
| +18 -86 -18 | 2099 | 0.002178 | 0.002333 | 0.000010 | 0.999239 | 0.000228 |

**Cluster +18 -86 -18**:
423 voxels (20%) covering 24% of atlas.LG r (Lingual Gyrus Right)
284 voxels (14%) covering 11% of atlas.Cereb1 r (Cerebelum Crus1 Right)
280 voxels (13%) covering 32% of atlas.OFusG r (Occipital Fusiform Gyrus Right)

252 voxels (12%) covering 32% of atlas.toITG r (Inferior Temporal Gyrus, temporooccipital part Right)
162 voxels (8%) covering 8% of atlas.iLOC r (Lateral Occipital Cortex, inferior division Right)
137 voxels (7%) covering 9% of atlas.Cereb6 r (Cerebelum 6 Right)
119 voxels (6%) covering 15% of atlas.TOFusC r (Temporal Occipital Fusiform Cortex Right)
107 voxels (5%) covering 4% of atlas.OP r (Occipital Pole Right)
25 voxels (1%) covering 2% of atlas.toMTG r (Middle Temporal Gyrus, temporooccipital part Right)
25 voxels (1%) covering 4% of atlas.Cereb45 r (Cerebelum 4 5 Right)
3 voxels (0%) covering 0% of atlas.pTFusC r (Temporal Fusiform Cortex, posterior division Right)
1 voxels (0%) covering 0% of atlas.pPaHC r (Parahippocampal Gyrus, posterior division Right)
1 voxels (0%) covering 0% of atlas.Ver45 (Vermis 4 5)
280 voxels (13%) covering 0% of atlas.not-labeled

**Table S13. SBC maps differences between aC and nC using POS2 as the seed.**

| Cluster (x,y,z) | size | size p-FWE | size p-FDR | size p-unc | peak p-FWE | peak p-unc |
|---|---|---|---|---|---|---|
| -14 +40 -06 | 4037 | 0.000002 | 0.000002 | 0.000000 | 0.934467 | 0.000062 |
| +30 +22 -12 | 1491 | 0.016131 | 0.007761 | 0.000070 | 1.000000 | 0.000568 |

**Cluster -14 +40 -6**:
1064 voxels (26%) covering 41% of atlas.AC (Cingulate Gyrus, anterior division)
829 voxels (21%) covering 10% of atlas.FP r (Frontal Pole Right)
319 voxels (8%) covering 12% of atlas.SFG r (Superior Frontal Gyrus Right)
182 voxels (5%) covering 13% of atlas.PaCiG r (Paracingulate Gyrus Right)
176 voxels (4%) covering 3% of atlas.FP l (Frontal Pole Left)
108 voxels (3%) covering 8% of atlas.PaCiG l (Paracingulate Gyrus Left)
105 voxels (3%) covering 11% of atlas.MedFC (Frontal Medial Cortex)
104 voxels (3%) covering 15% of atlas.SMA r (Juxtapositional Lobule Cortex -formerly Supplementary Motor Cortex- Right)
78 voxels (2%) covering 12% of atlas.SMA L(Juxtapositional Lobule Cortex -formerly Supplementary Motor Cortex- Left)
7 voxels (0%) covering 0% of atlas.FOrb r (Frontal Orbital Cortex Right)
3 voxels (0%) covering 0% of atlas.SubCalC (Subcallosal Cortex)
2 voxels (0%) covering 0% of atlas.MidFG r (Middle Frontal Gyrus Right)
1 voxels (0%) covering 0% of atlas.FOrb l (Frontal Orbital Cortex Left)
1059 voxels (26%) covering 0% of atlas.not-labeled

**Cluster +30 +22 -12**:
390 voxels (26%) covering 29% of atlas.IC r (Insular Cortex Right)
215 voxels (14%) covering 25% of atlas.CO r (Central Opercular Cortex Right)
168 voxels (11%) covering 7% of atlas.TP r (Temporal Pole Right)
157 voxels (11%) covering 11% of atlas.FOrb r (Frontal Orbital Cortex Right)
128 voxels (9%) covering 34% of atlas.PP r (Planum Polare Right)
69 voxels (5%) covering 2% of atlas.PreCG r (Precentral Gyrus Right)
45 voxels (3%) covering 14% of atlas.FO r (Frontal Operculum Cortex Right)
36 voxels (2%) covering 5% of atlas.IFG oper r (Inferior Frontal Gyrus, pars opercularis Right)
9 voxels (1%) covering 3% of atlas.aSTG r (Superior Temporal Gyrus, anterior division Right)
274 voxels (18%) covering 0% of atlas.not-labeled

**Table S14. SBC map association with MoCA using the whole Precuneus as seed.**

| Cluster (x,y,z) | Size (vox) | size p-FWE | size p-FDR | size p-unc | peak p-FWE | peak p-unc |
|---|---|---|---|---|---|---|
| +00 -70 +36 | 10496 | 0.000000 | 0.000000 | 0.000000 | 0.000404 | 0.000000 |
| -36 -50 -14 | 1542 | 0.002734 | 0.001389 | 0.000009 | 0.997704 | 0.000126 |
| +56 +38 +14 | 870 | 0.102299 | 0.036495 | 0.000367 | 1.000000 | 0.000978 |

**Cluster +0 -70 +36**:
4089 voxels (39%) covering 73% of atlas.Precuneous (Precuneous Cortex)
1566 voxels (15%) covering 65% of atlas.PC (Cingulate Gyrus, posterior division)
1207 voxels (11%) covering 25% of atlas.sLOC r (Lateral Occipital Cortex, superior division Right)
799 voxels (8%) covering 16% of atlas.sLOC l (Lateral Occipital Cortex, superior division Left)
475 voxels (5%) covering 32% of atlas.AG r (Angular Gyrus Right)
300 voxels (3%) covering 47% of atlas.Cuneal r (Cuneal Cortex Right)
171 voxels (2%) covering 33% of atlas.Cuneal l (Cuneal Cortex Left)
163 voxels (2%) covering 13% of atlas.pSMG r (Supramarginal Gyrus, posterior division Right)
81 voxels (1%) covering 11% of atlas.ICC r (Intracalcarine Cortex Right)
67 voxels (1%) covering 5% of atlas.SPL l (Superior Parietal Lobule Left)
47 voxels (0%) covering 2% of atlas.iLOC l (Lateral Occipital Cortex, inferior division Left)
34 voxels (0%) covering 2% of atlas.SPL r (Superior Parietal Lobule Right)
22 voxels (0%) covering 1% of atlas.PreCG l (Precentral Gyrus Left)
14 voxels (0%) covering 1% of atlas.AC (Cingulate Gyrus, anterior division)
13 voxels (0%) covering 0% of atlas.PreCG r (Precentral Gyrus Right)
10 voxels (0%) covering 1% of atlas.AG l (Angular Gyrus Left)
9 voxels (0%) covering 1% of atlas.toMTG r (Middle Temporal Gyrus, temporooccipital part Right)
8 voxels (0%) covering 1% of atlas.ICC l (Intracalcarine Cortex Left)
7 voxels (0%) covering 5% of atlas.SCC r (Supracalcarine Cortex Right)
2 voxels (0%) covering 0% of atlas.PostCG r (Postcentral Gyrus Right)
1 voxels (0%) covering 1% of atlas.SCC l (Supracalcarine Cortex Left)
1411 voxels (13%) covering 0% of atlas.not-labeled

**Cluster -36 -50 -14**:
306 voxels (20%) covering 40% of atlas.Hippocampus l
174 voxels (11%) covering 27% of atlas.TOFusC l (Temporal Occipital Fusiform Cortex Left)
141 voxels (9%) covering 7% of atlas.iLOC l (Lateral Occipital Cortex, inferior division Left)
137 voxels (9%) covering 35% of atlas.pPaHC l (Parahippocampal Gyrus, posterior division Left)
127 voxels (8%) covering 5% of atlas.OP l (Occipital Pole Left)
74 voxels (5%) covering 8% of atlas.OFusG l (Occipital Fusiform Gyrus Left)
64 voxels (4%) covering 20% of atlas.Amygdala l
39 voxels (3%) covering 5% of atlas.pTFusC l (Temporal Fusiform Cortex, posterior division Left)
31 voxels (2%) covering 1% of atlas.sLOC l (Lateral Occipital Cortex, superior division Left)
22 voxels (1%) covering 4% of atlas.aPaHC l (Parahippocampal Gyrus, anterior division Left)
18 voxels (1%) covering 3% of atlas.toITG l (Inferior Temporal Gyrus, temporooccipital part Left)

16 voxels (1%) covering 0% of atlas.Brain-Stem
13 voxels (1%) covering 2% of atlas.ICC l (Intracalcarine Cortex Left)
2 voxels (0%) covering 0% of atlas.Cereb6 l (Cerebelum 6 Left)
378 voxels (25%) covering 0% of atlas.not-labeled

**Cluster +56 +38 +14**:
270 voxels (31%) covering 10% of atlas.MidFG r (Middle Frontal Gyrus Right)
90 voxels (10%) covering 3% of atlas.AC (Cingulate Gyrus, anterior division)
84 voxels (10%) covering 6% of atlas.PaCiG r (Paracingulate Gyrus Right)
48 voxels (6%) covering 1% of atlas.FP r (Frontal Pole Right)
23 voxels (3%) covering 1% of atlas.SFG r (Superior Frontal Gyrus Right)
23 voxels (3%) covering 4% of atlas.IFG tri r (Inferior Frontal Gyrus, pars triangularis Right)
1 voxels (0%) covering 0% of atlas.PaCiG l (Paracingulate Gyrus Left)
331 voxels (38%) covering 0% of atlas.not-labeled

**Table S15. SBC map association with MoCA using 7Pm as seed.**

| Cluster (x,y,z) | Size (vox) | size p-FWE | size p-FDR | size p-unc | peak p-FWE | peak p-unc |
|---|---|---|---|---|---|---|
| -06 -66 +50 | 4039 | 0.000000 | 0.000000 | 0.000000 | 0.000003 | 0.000000 |
| -26 +22 -22 | 1149 | 0.023423 | 0.010373 | 0.000082 | 0.913607 | 0.000041 |
| +40 +08 -40 | 1073 | 0.035417 | 0.010520 | 0.000125 | 0.999689 | 0.000185 |
| -28 -14 -22 | 887 | 0.099923 | 0.023036 | 0.000364 | 0.916769 | 0.000042 |

**Cluster -6 -66 +50**:
2216 voxels (55%) covering 39% of atlas.Precuneous (Precuneous Cortex)
420 voxels (10%) covering 18% of atlas.PC (Cingulate Gyrus, posterior division)
324 voxels (8%) covering 7% of atlas.sLOC r (Lateral Occipital Cortex, superior division Right)
250 voxels (6%) covering 5% of atlas.sLOC l (Lateral Occipital Cortex, superior division Left)
57 voxels (1%) covering 4% of atlas.SPL r (Superior Parietal Lobule Right)
29 voxels (1%) covering 2% of atlas.SPL l (Superior Parietal Lobule Left)
5 voxels (0%) covering 0% of atlas.PreCG l (Precentral Gyrus Left)
5 voxels (0%) covering 0% of atlas.AC (Cingulate Gyrus, anterior division)
2 voxels (0%) covering 0% of atlas.PreCG r (Precentral Gyrus Right)
731 voxels (18%) covering 0% of atlas.not-labeled

**Cluster -26 +22 -22**:
402 voxels (35%) covering 17% of atlas.TP l (Temporal Pole Left)
253 voxels (22%) covering 15% of atlas.FOrb l (Frontal Orbital Cortex Left)
157 voxels (14%) covering 20% of atlas.IFG oper l (Inferior Frontal Gyrus, pars opercularis Left)
107 voxels (9%) covering 16% of atlas.IFG tri l (Inferior Frontal Gyrus, pars triangularis Left)
29 voxels (3%) covering 8% of atlas.FO l (Frontal Operculum Cortex Left)
8 voxels (1%) covering 1% of atlas.IC l (Insular Cortex Left)
8 voxels (1%) covering 1% of atlas.CO l (Central Opercular Cortex Left)
6 voxels (1%) covering 2% of atlas.PP l (Planum Polare Left)

3 voxels (0%) covering 0% of atlas.PreCG l (Precentral Gyrus Left)
3 voxels (0%) covering 1% of atlas.aSTG l (Superior Temporal Gyrus, anterior division Left)
173 voxels (15%) covering 0% of atlas.not-labeled

**Cluster +40 +8 -40**:
502 voxels (47%) covering 21% of atlas.TP r (Temporal Pole Right)
177 voxels (16%) covering 54% of atlas.aITG r (Inferior Temporal Gyrus, anterior division Right)
114 voxels (11%) covering 28% of atlas.aMTG r (Middle Temporal Gyrus, anterior division Right)
45 voxels (4%) covering 5% of atlas.pITG r (Inferior Temporal Gyrus, posterior division Right)
24 voxels (2%) covering 2% of atlas.pMTG r (Middle Temporal Gyrus, posterior division Right)
17 voxels (2%) covering 6% of atlas.aSTG r (Superior Temporal Gyrus, anterior division Right)
1 voxels (0%) covering 0% of atlas.pTFusC r (Temporal Fusiform Cortex, posterior division Right)
193 voxels (18%) covering 0% of atlas.not-labeled

**Cluster -28 -14 -22**:
371 voxels (42%) covering 49% of atlas.Hippocampus l
105 voxels (12%) covering 27% of atlas.pPaHC l (Parahippocampal Gyrus, posterior division Left)
76 voxels (9%) covering 13% of atlas.aPaHC l (Parahippocampal Gyrus, anterior division Left)
63 voxels (7%) covering 7% of atlas.pTFusC l (Temporal Fusiform Cortex, posterior division Left)
52 voxels (6%) covering 16% of atlas.Amygdala l
20 voxels (2%) covering 1% of atlas.TP l (Temporal Pole Left)
2 voxels (0%) covering 0% of atlas.pITG l (Inferior Temporal Gyrus, posterior division Left)
2 voxels (0%) covering 0% of atlas.Brain-Stem
1 voxels (0%) covering 0% of atlas.FOrb l (Frontal Orbital Cortex Left)
195 voxels (22%) covering 0% of atlas.not-labeled

**Table S16. SBC map association with MoCA using 7Am as seed.**

| Cluster (x,y,z) | Size (vox) | size p-FWE | size p-FDR | size p-unc | peak p-FWE | peak p-unc |
|---|---|---|---|---|---|---|
| -08 -58 +62 | 5643 | 0.000000 | 0.000000 | 0.000000 | 0.000007 | 0.000000 |
| +28 +00 +60 | 1554 | 0.002808 | 0.001346 | 0.000010 | 0.999870 | 0.000209 |

**Cluster -8 -58 +62**:
2031 voxels (36%) covering 36% of atlas.Precuneous (Precuneous Cortex)
928 voxels (16%) covering 19% of atlas.sLOC r (Lateral Occipital Cortex, superior division Right)
856 voxels (15%) covering 17% of atlas.sLOC l (Lateral Occipital Cortex, superior division Left)
539 voxels (10%) covering 37% of atlas.SPL r (Superior Parietal Lobule Right)
203 voxels (4%) covering 14% of atlas.SPL l (Superior Parietal Lobule Left)
119 voxels (2%) covering 5% of atlas.PC (Cingulate Gyrus, posterior division)
104 voxels (2%) covering 3% of atlas.PostCG r (Postcentral Gyrus Right)
58 voxels (1%) covering 2% of atlas.PostCG l (Postcentral Gyrus Left)
32 voxels (1%) covering 6% of atlas.Cuneal l (Cuneal Cortex Left)
24 voxels (0%) covering 1% of atlas.PreCG l (Precentral Gyrus Left)
20 voxels (0%) covering 1% of atlas.AG r (Angular Gyrus Right)

14 voxels (0%) covering 1% of atlas.pSMG r (Supramarginal Gyrus, posterior division Right)
11 voxels (0%) covering 0% of atlas.PreCG r (Precentral Gyrus Right)
10 voxels (0%) covering 2% of atlas.Cuneal r (Cuneal Cortex Right)
694 voxels (12%) covering 0% of atlas.not-labeled

**Cluster +28 +0 +60**:
269 voxels (17%) covering 10% of atlas.SFG r (Superior Frontal Gyrus Right)
177 voxels (11%) covering 13% of atlas.PaCiG l (Paracingulate Gyrus Left)
150 voxels (10%) covering 5% of atlas.MidFG r (Middle Frontal Gyrus Right)
148 voxels (10%) covering 5% of atlas.MidFG l (Middle Frontal Gyrus Left)
104 voxels (7%) covering 16% of atlas.SMA L(Juxtapositional Lobule Cortex -formerly Supplementary Motor Cortex- Left)
88 voxels (6%) covering 3% of atlas.AC (Cingulate Gyrus, anterior division)
71 voxels (5%) covering 10% of atlas.SMA r (Juxtapositional Lobule Cortex -formerly Supplementary Motor Cortex- Right)
47 voxels (3%) covering 1% of atlas.FP l (Frontal Pole Left)
38 voxels (2%) covering 3% of atlas.PaCiG r (Paracingulate Gyrus Right)
37 voxels (2%) covering 1% of atlas.SFG l (Superior Frontal Gyrus Left)
28 voxels (2%) covering 1% of atlas.PreCG r (Precentral Gyrus Right)
397 voxels (26%) covering 0% of atlas.not-labeled

**Table S17. SBC map association with MoCA using 7m as seed.**

| Cluster (x,y,z) | Size (vox) | size p-FWE | size p-FDR | size p-unc | peak p-FWE | peak p-unc |
|---|---|---|---|---|---|---|
| +00 -62 +32 | 6532 | 0.000000 | 0.000000 | 0.000000 | 0.000022 | 0.000000 |
| +12 +52 +16 | 2538 | 0.000038 | 0.000018 | 0.000000 | 0.796364 | 0.000025 |
| +54 -56 +38 | 2204 | 0.000159 | 0.000049 | 0.000001 | 0.085394 | 0.000001 |
| +12 +32 +40 | 1520 | 0.003736 | 0.000861 | 0.000013 | 0.382238 | 0.000006 |
| -46 -64 +34 | 1241 | 0.015393 | 0.002856 | 0.000054 | 0.716117 | 0.000018 |
| -48 +18 +50 | 827 | 0.147133 | 0.024417 | 0.000557 | 0.819772 | 0.000027 |

**Cluster +0 -62 +32**:
2923 voxels (45%) covering 52% of atlas.Precuneous (Precuneous Cortex)
1555 voxels (24%) covering 65% of atlas.PC (Cingulate Gyrus, posterior division)
240 voxels (4%) covering 37% of atlas.Cuneal r (Cuneal Cortex Right)
158 voxels (2%) covering 30% of atlas.Cuneal l (Cuneal Cortex Left)
45 voxels (1%) covering 1% of atlas.sLOC r (Lateral Occipital Cortex, superior division Right)
39 voxels (1%) covering 7% of atlas.Caudate l
13 voxels (0%) covering 0% of atlas.sLOC l (Lateral Occipital Cortex, superior division Left)
12 voxels (0%) covering 0% of atlas.AC (Cingulate Gyrus, anterior division)
6 voxels (0%) covering 4% of atlas.SCC r (Supracalcarine Cortex Right)
5 voxels (0%) covering 0% of atlas.Thalamus l
1 voxels (0%) covering 1% of atlas.SCC l (Supracalcarine Cortex Left)
1 voxels (0%) covering 0% of atlas.Caudate r

1534 voxels (23%) covering 0% of atlas.not-labeled

**Cluster +12 +52 +16**:
1254 voxels (49%) covering 16% of atlas.FP r (Frontal Pole Right)
283 voxels (11%) covering 21% of atlas.PaCiG r (Paracingulate Gyrus Right)
256 voxels (10%) covering 26% of atlas.MedFC (Frontal Medial Cortex)
127 voxels (5%) covering 2% of atlas.FP l (Frontal Pole Left)
56 voxels (2%) covering 2% of atlas.SFG r (Superior Frontal Gyrus Right)
55 voxels (2%) covering 4% of atlas.PaCiG l (Paracingulate Gyrus Left)
45 voxels (2%) covering 2% of atlas.SFG l (Superior Frontal Gyrus Left)
16 voxels (1%) covering 1% of atlas.SubCalC (Subcallosal Cortex)
14 voxels (1%) covering 1% of atlas.FOrb r (Frontal Orbital Cortex Right)
12 voxels (0%) covering 0% of atlas.AC (Cingulate Gyrus, anterior division)
420 voxels (17%) covering 0% of atlas.not-labeled

**Cluster +54 -56 +38**:
987 voxels (45%) covering 21% of atlas.sLOC r (Lateral Occipital Cortex, superior division Right)
925 voxels (42%) covering 63% of atlas.AG r (Angular Gyrus Right)
54 voxels (2%) covering 4% of atlas.pSMG r (Supramarginal Gyrus, posterior division Right)
238 voxels (11%) covering 0% of atlas.not-labeled

**Cluster +12 +32 +40**:
294 voxels (19%) covering 11% of atlas.SFG r (Superior Frontal Gyrus Right)
253 voxels (17%) covering 9% of atlas.SFG l (Superior Frontal Gyrus Left)
171 voxels (11%) covering 13% of atlas.PaCiG r (Paracingulate Gyrus Right)
156 voxels (10%) covering 2% of atlas.FP r (Frontal Pole Right)
147 voxels (10%) covering 2% of atlas.FP l (Frontal Pole Left)
47 voxels (3%) covering 2% of atlas.MidFG r (Middle Frontal Gyrus Right)
28 voxels (2%) covering 1% of atlas.MidFG l (Middle Frontal Gyrus Left)
21 voxels (1%) covering 2% of atlas.PaCiG l (Paracingulate Gyrus Left)
403 voxels (27%) covering 0% of atlas.not-labeled

**Cluster -46 -64 +34**:
789 voxels (64%) covering 16% of atlas.sLOC l (Lateral Occipital Cortex, superior division Left)
371 voxels (30%) covering 39% of atlas.AG l (Angular Gyrus Left)
57 voxels (5%) covering 5% of atlas.pSMG l (Supramarginal Gyrus, posterior division Left)
4 voxels (0%) covering 0% of atlas.iLOC l (Lateral Occipital Cortex, inferior division Left)
20 voxels (2%) covering 0% of atlas.not-labeled

**Cluster -48 +18 +50**:
581 voxels (70%) covering 20% of atlas.MidFG l (Middle Frontal Gyrus Left)
59 voxels (7%) covering 9% of atlas.IFG tri l (Inferior Frontal Gyrus, pars triangularis Left)
35 voxels (4%) covering 5% of atlas.IFG oper l (Inferior Frontal Gyrus, pars opercularis Left)
2 voxels (0%) covering 0% of atlas.FP l (Frontal Pole Left)

150 voxels (18%) covering 0% of atlas.not-labeled

**Table S18. SBC map association with MoCA using PCV as seed.**

| Cluster (x,y,z) | Size (vox) | size p-FWE | size p-FDR | size p-unc | peak p-FWE | peak p-unc |
|---|---|---|---|---|---|---|
| +02 -50 +48 | 5244 | 0.000000 | 0.000000 | 0.000000 | 0.000005 | 0.000000 |
| +24 -30 -42 | 1248 | 0.014480 | 0.007347 | 0.000051 | 0.999999 | 0.000392 |

**Cluster +2 -50 +48**:
2420 voxels (46%) covering 43% of atlas.Precuneous (Precuneous Cortex)
680 voxels (13%) covering 28% of atlas.PC (Cingulate Gyrus, posterior division)
180 voxels (3%) covering 12% of atlas.SPL r (Superior Parietal Lobule Right)
134 voxels (3%) covering 3% of atlas.sLOC r (Lateral Occipital Cortex, superior division Right)
79 voxels (2%) covering 2% of atlas.PostCG l (Postcentral Gyrus Left)
76 voxels (1%) covering 12% of atlas.Cuneal r (Cuneal Cortex Right)
72 voxels (1%) covering 2% of atlas.PostCG r (Postcentral Gyrus Right)
61 voxels (1%) covering 2% of atlas.AC (Cingulate Gyrus, anterior division)
40 voxels (1%) covering 1% of atlas.sLOC l (Lateral Occipital Cortex, superior division Left)
34 voxels (1%) covering 1% of atlas.PreCG l (Precentral Gyrus Left)
32 voxels (1%) covering 2% of atlas.SPL l (Superior Parietal Lobule Left)
20 voxels (0%) covering 3% of atlas.SMA L(Juxtapositional Lobule Cortex -formerly Supplementary Motor Cortex- Left)
5 voxels (0%) covering 0% of atlas.PreCG r (Precentral Gyrus Right)
3 voxels (0%) covering 0% of atlas.ICC r (Intracalcarine Cortex Right)
2 voxels (0%) covering 1% of atlas.SCC r (Supracalcarine Cortex Right)
1406 voxels (27%) covering 0% of atlas.not-labeled

**Cluster +24 -30 -42**:
345 voxels (28%) covering 8% of atlas.Brain-Stem
139 voxels (11%) covering 15% of atlas.pITG r (Inferior Temporal Gyrus, posterior division Right)
78 voxels (6%) covering 3% of atlas.Cereb1 r (Cerebelum Crus1 Right)
71 voxels (6%) covering 11% of atlas.aPaHC r (Parahippocampal Gyrus, anterior division Right)
55 voxels (4%) covering 34% of atlas.Cereb10 r (Cerebelum 10 Right)
42 voxels (3%) covering 6% of atlas.pTFusC r (Temporal Fusiform Cortex, posterior division Right)
30 voxels (2%) covering 5% of atlas.aPaHC l (Parahippocampal Gyrus, anterior division Left)
25 voxels (2%) covering 1% of atlas.Cereb8 r (Cerebelum 8 Right)
25 voxels (2%) covering 17% of atlas.Cereb10 l (Cerebelum 10 Left)
21 voxels (2%) covering 3% of atlas.Cereb45 r (Cerebelum 4 5 Right)
20 voxels (2%) covering 1% of atlas.Cereb6 r (Cerebelum 6 Right)
17 voxels (1%) covering 1% of atlas.pMTG r (Middle Temporal Gyrus, posterior division Right)
8 voxels (1%) covering 1% of atlas.pTFusC l (Temporal Fusiform Cortex, posterior division Left)
6 voxels (0%) covering 1% of atlas.TOFusC r (Temporal Occipital Fusiform Cortex Right)
3 voxels (0%) covering 0% of atlas.Hippocampus r
2 voxels (0%) covering 1% of atlas.Amygdala r

1 voxels (0%) covering 0% of atlas.aTFusC r (Temporal Fusiform Cortex, anterior division Right)
1 voxels (0%) covering 0% of atlas.Cereb2 r (Cerebelum Crus2 Right)
1 voxels (0%) covering 0% of atlas.Cereb7 r (Cerebelum 7b Right)
358 voxels (29%) covering 0% of atlas.not-labeled

**Table S19. SBC map association with MoCA using POS2 as seed.**

| Cluster (x,y,z) | Size (vox) | size p-FWE | size p-FDR | size p-unc | peak p-FWE | peak p-unc |
|---|---|---|---|---|---|---|
| -04 -70 +36 | 6897 | 0.000000 | 0.000000 | 0.000000 | 0.000095 | 0.000000 |
| -36 -50 -14 | 1452 | 0.004649 | 0.002317 | 0.0000016 | 0.977684 | 0.000071 |
| +28 +68 +02 | 1245 | 0.013575 | 0.004531 | 0.000047 | 0.999722 | 0.000186 |
| -24 +68 +06 | 818 | 0.144285 | 0.033992 | 0.000536 | 0.047907 | 0.000000 |
| +34 -28 -12 | 803 | 0.157089 | 0.033992 | 0.000588 | 0.999986 | 0.000279 |

**Cluster -4 -70 +36**:
2881 voxels (42%) covering 51% of atlas.Precuneous (Precuneous Cortex)
1099 voxels (16%) covering 46% of atlas.PC (Cingulate Gyrus, posterior division)
711 voxels (10%) covering 15% of atlas.sLOC r (Lateral Occipital Cortex, superior division Right)
289 voxels (4%) covering 45% of atlas.Cuneal r (Cuneal Cortex Right)
223 voxels (3%) covering 4% of atlas.sLOC l (Lateral Occipital Cortex, superior division Left)
201 voxels (3%) covering 14% of atlas.AG r (Angular Gyrus Right)
177 voxels (3%) covering 34% of atlas.Cuneal l (Cuneal Cortex Left)
66 voxels (1%) covering 5% of atlas.pSMG r (Supramarginal Gyrus, posterior division Right)
57 voxels (1%) covering 4% of atlas.SPL l (Superior Parietal Lobule Left)
24 voxels (0%) covering 1% of atlas.PreCG r (Precentral Gyrus Right)
7 voxels (0%) covering 2% of atlas.PT r (Planum Temporale Right)
2 voxels (0%) covering 0% of atlas.AC (Cingulate Gyrus, anterior division)
1 voxels (0%) covering 0% of atlas.PostCG r (Postcentral Gyrus Right)
1159 voxels (17%) covering 0% of atlas.not-labeled

**Cluster -36 -50 -14**:
221 voxels (15%) covering 29% of atlas.Hippocampus l
198 voxels (14%) covering 51% of atlas.pPaHC l (Parahippocampal Gyrus, posterior division Left)
132 voxels (9%) covering 6% of atlas.iLOC l (Lateral Occipital Cortex, inferior division Left)
123 voxels (8%) covering 19% of atlas.TOFusC l (Temporal Occipital Fusiform Cortex Left)
69 voxels (5%) covering 21% of atlas.Amygdala l
61 voxels (4%) covering 7% of atlas.pTFusC l (Temporal Fusiform Cortex, posterior division Left)
44 voxels (3%) covering 6% of atlas.toITG l (Inferior Temporal Gyrus, temporooccipital part Left)
41 voxels (3%) covering 1% of atlas.Brain-Stem
16 voxels (1%) covering 2% of atlas.Cereb45 l (Cerebelum 4 5 Left)
14 voxels (1%) covering 2% of atlas.OFusG l (Occipital Fusiform Gyrus Left)
3 voxels (0%) covering 1% of atlas.aPaHC l (Parahippocampal Gyrus, anterior division Left)
2 voxels (0%) covering 0% of atlas.Thalamus l
1 voxels (0%) covering 0% of atlas.toMTG l (Middle Temporal Gyrus, temporooccipital part Left)

1 voxels (0%) covering 0% of atlas.pITG l (Inferior Temporal Gyrus, posterior division Left)
1 voxels (0%) covering 1% of atlas.Cereb3 l (Cerebelum 3 Left)
525 voxels (36%) covering 0% of atlas.not-labeled

**Cluster +28 +68 +2**:
655 voxels (53%) covering 8% of atlas.FP r (Frontal Pole Right)
222 voxels (18%) covering 16% of atlas.PaCiG r (Paracingulate Gyrus Right)
213 voxels (17%) covering 8% of atlas.AC (Cingulate Gyrus, anterior division)
26 voxels (2%) covering 2% of atlas.PaCiG l (Paracingulate Gyrus Left)
1 voxels (0%) covering 0% of atlas.SMA r (Juxtapositional Lobule Cortex -formerly Supplementary Motor Cortex- Right)
128 voxels (10%) covering 0% of atlas.not-labeled

**Cluster -24 +68 +6:**
751 voxels (92%) covering 11% of atlas.FP l (Frontal Pole Left)
67 voxels (8%) covering 0% of atlas.not-labeled

**Cluster +34 -28 -12**:
138 voxels (17%) covering 20% of atlas.Hippocampus r
118 voxels (15%) covering 37% of atlas.pPaHC r (Parahippocampal Gyrus, posterior division Right)
73 voxels (9%) covering 9% of atlas.TOFusC r (Temporal Occipital Fusiform Cortex Right)
53 voxels (7%) covering 7% of atlas.pTFusC r (Temporal Fusiform Cortex, posterior division Right)
38 voxels (5%) covering 6% of atlas.aPaHC r (Parahippocampal Gyrus, anterior division Right)
35 voxels (4%) covering 3% of atlas.Thalamus r
27 voxels (3%) covering 1% of atlas.Brain-Stem
17 voxels (2%) covering 7% of atlas.Ver3 (Vermis 3)
8 voxels (1%) covering 1% of atlas.pITG r (Inferior Temporal Gyrus, posterior division Right)
7 voxels (1%) covering 1% of atlas.toITG r (Inferior Temporal Gyrus, temporooccipital part Right)
3 voxels (0%) covering 2% of atlas.Cereb3 r (Cerebelum 3 Right)
2 voxels (0%) covering 1% of atlas.Pallidum r
1 voxels (0%) covering 0% of atlas.PC (Cingulate Gyrus, posterior division)
1 voxels (0%) covering 0% of atlas.Thalamus l
1 voxels (0%) covering 0% of atlas.Cereb45 r (Cerebelum 4 5 Right)
281 voxels (35%) covering 0% of atlas.not-labeled

**Table S20. SBC map association with CERAD's List of words using the whole Precuneus as seed.**

| Cluster (x,y,z) | Size (vox) | size p-FWE | size p-FDR | size p-unc | peak p-FWE | peak p-unc |
|---|---|---|---|---|---|---|
| +10 -70 +36 | 6535 | 0.000000 | 0.000000 | 0.000000 | 0.005831 | 0.000000 |
| +42 -70 +34 | 2082 | 0.000216 | 0.000105 | 0.000001 | 0.399047 | 0.000006 |

**Cluster +10 -70 +36** :
3490 voxels (53%) covering 62% of atlas.Precuneous (Precuneous Cortex)
1570 voxels (24%) covering 65% of atlas.PC (Cingulate Gyrus, posterior division)

236 voxels (4%) covering 37% of atlas.Cuneal r (Cuneal Cortex Right)
124 voxels (2%) covering 5% of atlas.AC (Cingulate Gyrus, anterior division)
79 voxels (1%) covering 15% of atlas.Cuneal l (Cuneal Cortex Left)
67 voxels (1%) covering 1% of atlas.sLOC r (Lateral Occipital Cortex, superior division Right)
53 voxels (1%) covering 1% of atlas.PreCG l (Precentral Gyrus Left)
32 voxels (0%) covering 1% of atlas.sLOC l (Lateral Occipital Cortex, superior division Left)
26 voxels (0%) covering 1% of atlas.PreCG r (Precentral Gyrus Right)
25 voxels (0%) covering 4% of atlas.SMA L(Juxtapositional Lobule Cortex -formerly Supplementary Motor Cortex- Left)
21 voxels (0%) covering 15% of atlas.SCC r (Supracalcarine Cortex Right)
19 voxels (0%) covering 1% of atlas.SPL l (Superior Parietal Lobule Left)
8 voxels (0%) covering 1% of atlas.ICC r (Intracalcarine Cortex Right)
7 voxels (0%) covering 10% of atlas.SCC l (Supracalcarine Cortex Left)
778 voxels (12%) covering 0% of atlas.not-labeled

**Cluster +42 -70 +34** :
668 voxels (32%) covering 14% of atlas.sLOC r (Lateral Occipital Cortex, superior division Right)
640 voxels (31%) covering 44% of atlas.AG r (Angular Gyrus Right)
203 voxels (10%) covering 16% of atlas.pSMG r (Supramarginal Gyrus, posterior division Right)
99 voxels (5%) covering 24% of atlas.pSTG r (Superior Temporal Gyrus, posterior division Right)
60 voxels (3%) covering 4% of atlas.pMTG r (Middle Temporal Gyrus, posterior division Right)
50 voxels (2%) covering 11% of atlas.PT r (Planum Temporale Right)
5 voxels (0%) covering 0% of atlas.toMTG r (Middle Temporal Gyrus, temporooccipital part Right)
1 voxels (0%) covering 0% of atlas.aMTG r (Middle Temporal Gyrus, anterior division Right)
1 voxels (0%) covering 0% of atlas.HG r (Heschl's Gyrus Right)
355 voxels (17%) covering 0% of atlas.not-labeled

**Table S21. SBC map association with CERAD's List of words using 7Pm as seed.**

| Cluster (x,y,z) | Size (vox) | size p-FWE | size p-FDR | size p-unc | peak p-FWE | peak p-unc |
|---|---|---|---|---|---|---|
| -02 -66 +48 | 1764 | 0.001087 | 0.001046 | 0.000004 | 0.493648 | 0.000009 |
| +22 -42 -06 | 1057 | 0.039466 | 0.019366 | 0.000140 | 0.780846 | 0.000023 |
| -30 -40 -08 | 892 | 0.098832 | 0.033366 | 0.000361 | 0.999422 | 0.000167 |

**Cluster -2 -66 +48**:
1169 voxels (66%) covering 21% of atlas.Precuneous (Precuneous Cortex)
359 voxels (20%) covering 7% of atlas.sLOC r (Lateral Occipital Cortex, superior division Right)
84 voxels (5%) covering 4% of atlas.PC (Cingulate Gyrus, posterior division)
20 voxels (1%) covering 1% of atlas.AG r (Angular Gyrus Right)
18 voxels (1%) covering 0% of atlas.PreCG l (Precentral Gyrus Left)
13 voxels (1%) covering 0% of atlas.sLOC l (Lateral Occipital Cortex, superior division Left)
101 voxels (6%) covering 0% of atlas.not-labeled

**Cluster +22 -42 -6**:
198 voxels (19%) covering 28% of atlas.Hippocampus r
155 voxels (15%) covering 9% of atlas.LG r (Lingual Gyrus Right)
129 voxels (12%) covering 40% of atlas.pPaHC r (Parahippocampal Gyrus, posterior division Right)
102 voxels (10%) covering 16% of atlas.aPaHC r (Parahippocampal Gyrus, anterior division Right)

44 voxels (4%) covering 7% of atlas.Ver45 (Vermis 4 5)
37 voxels (4%) covering 1% of atlas.Brain-Stem
29 voxels (3%) covering 5% of atlas.Cereb45 r (Cerebelum 4 5 Right)
28 voxels (3%) covering 3% of atlas.TOFusC r (Temporal Occipital Fusiform Cortex Right)
21 voxels (2%) covering 3% of atlas.pTFusC r (Temporal Fusiform Cortex, posterior division Right)
11 voxels (1%) covering 0% of atlas.PC (Cingulate Gyrus, posterior division)
9 voxels (1%) covering 4% of atlas.Ver3 (Vermis 3)
8 voxels (1%) covering 2% of atlas.Amygdala r
2 voxels (0%) covering 1% of atlas.Cereb3 r (Cerebelum 3 Right)
284 voxels (27%) covering 0% of atlas.not-labeled

**Cluster -30 -40 -8**:
192 voxels (22%) covering 25% of atlas.Hippocampus l
142 voxels (16%) covering 36% of atlas.pPaHC l (Parahippocampal Gyrus, posterior division Left)
96 voxels (11%) covering 16% of atlas.aPaHC l (Parahippocampal Gyrus, anterior division Left)
84 voxels (9%) covering 10% of atlas.pTFusC l (Temporal Fusiform Cortex, posterior division Left)
38 voxels (4%) covering 3% of atlas.LG l (Lingual Gyrus Left)
27 voxels (3%) covering 1% of atlas.TP l (Temporal Pole Left)
24 voxels (3%) covering 1% of atlas.Brain-Stem
20 voxels (2%) covering 6% of atlas.Amygdala l
14 voxels (2%) covering 2% of atlas.Cereb45 l (Cerebelum 4 5 Left)
7 voxels (1%) covering 0% of atlas.FOrb l (Frontal Orbital Cortex Left)
6 voxels (1%) covering 1% of atlas.TOFusC l (Temporal Occipital Fusiform Cortex Left)
3 voxels (0%) covering 2% of atlas.Cereb3 l (Cerebelum 3 Left)
239 voxels (27%) covering 0% of atlas.not-labeled

**Table S22. SBC map association with CERAD's List of words using 7Am as seed.**

| Cluster (x,y,z) | Size (vox) | size p-FWE | size p-FDR | size p-unc | peak p-FWE | peak p-unc |
|---|---|---|---|---|---|---|
| -10 -54 +62 | 1615 | 0.002176 | 0.002218 | 0.000008 | 0.761601 | 0.000021 |
| +06 -64 +08 | 1210 | 0.017028 | 0.008746 | 0.000059 | 1.000000 | 0.000445 |

**Cluster -10 -54 +62**:
910 voxels (56%) covering 16% of atlas.Precuneous (Precuneous Cortex)
202 voxels (13%) covering 14% of atlas.SPL l (Superior Parietal Lobule Left)
129 voxels (8%) covering 3% of atlas.sLOC l (Lateral Occipital Cortex, superior division Left)
112 voxels (7%) covering 2% of atlas.sLOC r (Lateral Occipital Cortex, superior division Right)
37 voxels (2%) covering 1% of atlas.PostCG l (Postcentral Gyrus Left)
28 voxels (2%) covering 2% of atlas.SPL r (Superior Parietal Lobule Right)
6 voxels (0%) covering 0% of atlas.PC (Cingulate Gyrus, posterior division)
2 voxels (0%) covering 0% of atlas.PreCG r (Precentral Gyrus Right)
189 voxels (12%) covering 0% of atlas.not-labeled

**Cluster +6 -64 +8**:

365 voxels (30%) covering 7% of atlas.Precuneous (Precuneous Cortex)
260 voxels (21%) covering 35% of atlas.ICC r (Intracalcarine Cortex Right)
234 voxels (19%) covering 37% of atlas.ICC l (Intracalcarine Cortex Left)
76 voxels (6%) covering 15% of atlas.Cuneal l (Cuneal Cortex Left)
62 voxels (5%) covering 4% of atlas.LG r (Lingual Gyrus Right)
53 voxels (4%) covering 3% of atlas.LG l (Lingual Gyrus Left)
18 voxels (1%) covering 13% of atlas.SCC r (Supracalcarine Cortex Right)
16 voxels (1%) covering 22% of atlas.SCC l (Supracalcarine Cortex Left)
1 voxels (0%) covering 0% of atlas.PC (Cingulate Gyrus, posterior division)
1 voxels (0%) covering 0% of atlas.Cuneal r (Cuneal Cortex Right)
124 voxels (10%) covering 0% of atlas.not-labeled

**Table S23. SBC map association with CERAD's List of words using 7m as seed.**

| Cluster (x,y,z) | Size (vox) | size p-FWE | size p-FDR | size p-unc | peak p-FWE | peak p-unc |
|---|---|---|---|---|---|---|
| +02 -62 +30 | 4983 | 0.000000 | 0.000000 | 0.000000 | 0.002523 | 0.000000 |
| +54 -54 +38 | 2115 | 0.000245 | 0.000116 | 0.000001 | 0.864921 | 0.000033 |
| -12 -94 -26 | 1117 | 0.030561 | 0.009623 | 0.000109 | 0.999998 | 0.000354 |
| -42 -68 +30 | 1067 | 0.040012 | 0.009623 | 0.000144 | 0.994198 | 0.000106 |
| +14 +48 +16 | 846 | 0.135126 | 0.027368 | 0.000511 | 0.997829 | 0.000133 |

**Cluster +2 -62 +30**:
2527 voxels (51%) covering 45% of atlas.Precuneous (Precuneous Cortex)
1413 voxels (28%) covering 59% of atlas.PC (Cingulate Gyrus, posterior division)
231 voxels (5%) covering 36% of atlas.Cuneal r (Cuneal Cortex Right)
32 voxels (1%) covering 1% of atlas.sLOC r (Lateral Occipital Cortex, superior division Right)
27 voxels (1%) covering 1% of atlas.PreCG l (Precentral Gyrus Left)
23 voxels (0%) covering 1% of atlas.PreCG r (Precentral Gyrus Right)
16 voxels (0%) covering 11% of atlas.SCC r (Supracalcarine Cortex Right)
8 voxels (0%) covering 0% of atlas.sLOC l (Lateral Occipital Cortex, superior division Left)
7 voxels (0%) covering 1% of atlas.Cuneal l (Cuneal Cortex Left)
7 voxels (0%) covering 10% of atlas.SCC l (Supracalcarine Cortex Left)
5 voxels (0%) covering 0% of atlas.SPL l (Superior Parietal Lobule Left)
4 voxels (0%) covering 0% of atlas.AC (Cingulate Gyrus, anterior division)
683 voxels (14%) covering 0% of atlas.not-labeled

**Cluster +54 -54 +38**:
937 voxels (44%) covering 64% of atlas.AG r (Angular Gyrus Right)
664 voxels (31%) covering 14% of atlas.sLOC r (Lateral Occipital Cortex, superior division Right)
252 voxels (12%) covering 20% of atlas.pSMG r (Supramarginal Gyrus, posterior division Right)
15 voxels (1%) covering 3% of atlas.PT r (Planum Temporale Right)
5 voxels (0%) covering 1% of atlas.PO r (Parietal Operculum Cortex Right)
242 voxels (11%) covering 0% of atlas.not-labeled

**Cluster -12 -94 -26**:
369 voxels (33%) covering 19% of atlas.Cereb2 l (Cerebelum Crus2 Left)
341 voxels (31%) covering 16% of atlas.Cereb2 r (Cerebelum Crus2 Right)
32 voxels (3%) covering 1% of atlas.Cereb1 r (Cerebelum Crus1 Right)
3 voxels (0%) covering 0% of atlas.Cereb1 l (Cerebelum Crus1 Left)
1 voxels (0%) covering 0% of atlas.Cereb7 l (Cerebelum 7b Left)
371 voxels (33%) covering 0% of atlas.not-labeled

**Cluster -42 -68 +30**:
583 voxels (55%) covering 12% of atlas.sLOC l (Lateral Occipital Cortex, superior division Left)
272 voxels (25%) covering 29% of atlas.AG l (Angular Gyrus Left)
75 voxels (7%) covering 7% of atlas.pSMG l (Supramarginal Gyrus, posterior division Left)
137 voxels (13%) covering 0% of atlas.not-labeled

**Cluster +14 +48 +16**:
305 voxels (36%) covering 4% of atlas.FP r (Frontal Pole Right)
115 voxels (14%) covering 4% of atlas.SFG r (Superior Frontal Gyrus Right)
91 voxels (11%) covering 7% of atlas.PaCiG r (Paracingulate Gyrus Right)
51 voxels (6%) covering 1% of atlas.FP l (Frontal Pole Left)
41 voxels (5%) covering 1% of atlas.SFG l (Superior Frontal Gyrus Left)
30 voxels (4%) covering 2% of atlas.PaCiG l (Paracingulate Gyrus Left)
1 voxels (0%) covering 0% of atlas.AC (Cingulate Gyrus, anterior division)
212 voxels (25%) covering 0% of atlas.not-labeled

**Table S24. SBC map association with CERAD's List of words using PCV as seed.**

| Cluster (x,y,z) | Size (vox) | size p-FWE | size p-FDR | size p-unc | peak p-FWE | peak p-unc |
|---|---|---|---|---|---|---|
| +10 -48 +44 | 4910 | 0.000000 | 0.000000 | 0.000000 | 0.029417 | 0.000000 |
| +08 +40 -12 | 1326 | 0.009823 | 0.005141 | 0.000035 | 0.999027 | 0.000154 |

**Cluster +10 -48 +44**:
2933 voxels (60%) covering 52% of atlas.Precuneous (Precuneous Cortex)
781 voxels (16%) covering 33% of atlas.PC (Cingulate Gyrus, posterior division)
99 voxels (2%) covering 4% of atlas.AC (Cingulate Gyrus, anterior division)
91 voxels (2%) covering 14% of atlas.Cuneal r (Cuneal Cortex Right)
68 voxels (1%) covering 9% of atlas.ICC r (Intracalcarine Cortex Right)
53 voxels (1%) covering 1% of atlas.PreCG l (Precentral Gyrus Left)
43 voxels (1%) covering 7% of atlas.ICC l (Intracalcarine Cortex Left)
40 voxels (1%) covering 28% of atlas.SCC r (Supracalcarine Cortex Right)
38 voxels (1%) covering 6% of atlas.SMA L(Juxtapositional Lobule Cortex -formerly Supplementary Motor Cortex- Left)
37 voxels (1%) covering 7% of atlas.Cuneal l (Cuneal Cortex Left)
16 voxels (0%) covering 22% of atlas.SCC l (Supracalcarine Cortex Left)
14 voxels (0%) covering 1% of atlas.LG r (Lingual Gyrus Right)

10 voxels (0%) covering 2% of atlas.Ver45 (Vermis 4 5)
7 voxels (0%) covering 0% of atlas.sLOC r (Lateral Occipital Cortex, superior division Right)
7 voxels (0%) covering 0% of atlas.LG l (Lingual Gyrus Left)
3 voxels (0%) covering 0% of atlas.PreCG r (Precentral Gyrus Right)
1 voxels (0%) covering 0% of atlas.PostCG l (Postcentral Gyrus Left)
1 voxels (0%) covering 0% of atlas.SPL r (Superior Parietal Lobule Right)
1 voxels (0%) covering 0% of atlas.Thalamus l
1 voxels (0%) covering 0% of atlas.Cereb45 l (Cerebelum 4 5 Left)
666 voxels (14%) covering 0% of atlas.not-labeled

**Cluster +8 +40 -12**:
326 voxels (25%) covering 33% of atlas.MedFC (Frontal Medial Cortex)
215 voxels (16%) covering 3% of atlas.FP l (Frontal Pole Left)
190 voxels (14%) covering 17% of atlas.SubCalC (Subcallosal Cortex)
138 voxels (10%) covering 2% of atlas.FP r (Frontal Pole Right)
109 voxels (8%) covering 8% of atlas.PaCiG l (Paracingulate Gyrus Left)
17 voxels (1%) covering 1% of atlas.PaCiG r (Paracingulate Gyrus Right)
10 voxels (1%) covering 0% of atlas.AC (Cingulate Gyrus, anterior division)
5 voxels (0%) covering 0% of atlas.FOrb r (Frontal Orbital Cortex Right)
316 voxels (24%) covering 0% of atlas.not-labeled

**Table S25. SBC map association with CERAD's List of words using POS2 as seed.**

| Cluster (x,y,z) | Size (vox) | size p-FWE | size p-FDR | size p-unc | peak p-FWE | peak p-unc |
|---|---|---|---|---|---|---|
| -02 -74 +32 | 5703 | 0.000000 | 0.000000 | 0.000000 | 0.000617 | 0.000000 |
| +40 -68 +38 | 1189 | 0.018514 | 0.006224 | 0.000064 | 0.999995 | 0.000311 |
| +14 -32 -10 | 1182 | 0.019222 | 0.006224 | 0.000067 | 0.944227 | 0.000050 |

**Cluster -2 -74 +32**:
2675 voxels (47%) covering 48% of atlas.Precuneous (Precuneous Cortex)
1465 voxels (26%) covering 61% of atlas.PC (Cingulate Gyrus, posterior division)
264 voxels (5%) covering 41% of atlas.Cuneal r (Cuneal Cortex Right)
152 voxels (3%) covering 6% of atlas.AC (Cingulate Gyrus, anterior division)
98 voxels (2%) covering 19% of atlas.Cuneal l (Cuneal Cortex Left)
59 voxels (1%) covering 1% of atlas.PreCG l (Precentral Gyrus Left)
59 voxels (1%) covering 1% of atlas.sLOC r (Lateral Occipital Cortex, superior division Right)
55 voxels (1%) covering 1% of atlas.sLOC l (Lateral Occipital Cortex, superior division Left)
40 voxels (1%) covering 1% of atlas.PreCG r (Precentral Gyrus Right)
13 voxels (0%) covering 9% of atlas.SCC r (Supracalcarine Cortex Right)
9 voxels (0%) covering 1% of atlas.SPL l (Superior Parietal Lobule Left)
2 voxels (0%) covering 3% of atlas.SCC l (Supracalcarine Cortex Left)
1 voxels (0%) covering 0% of atlas.PostCG r (Postcentral Gyrus Right)
811 voxels (14%) covering 0% of atlas.not-labeled

**Cluster +40 -68 +38**:
284 voxels (24%) covering 19% of atlas.AG r (Angular Gyrus Right)
248 voxels (21%) covering 5% of atlas.sLOC r (Lateral Occipital Cortex, superior division Right)
177 voxels (15%) covering 14% of atlas.pSMG r (Supramarginal Gyrus, posterior division Right)
85 voxels (7%) covering 20% of atlas.pSTG r (Superior Temporal Gyrus, posterior division Right)
48 voxels (4%) covering 4% of atlas.pMTG r (Middle Temporal Gyrus, posterior division Right)
32 voxels (3%) covering 7% of atlas.PT r (Planum Temporale Right)
5 voxels (0%) covering 0% of atlas.toMTG r (Middle Temporal Gyrus, temporooccipital part Right)
4 voxels (0%) covering 1% of atlas.aMTG r (Middle Temporal Gyrus, anterior division Right)
306 voxels (26%) covering 0% of atlas.not-labeled

**Cluster +14 -32 -10**:
181 voxels (15%) covering 24% of atlas.Hippocampus l
152 voxels (13%) covering 26% of atlas.aPaHC l (Parahippocampal Gyrus, anterior division Left)
148 voxels (13%) covering 6% of atlas.TP l (Temporal Pole Left)
74 voxels (6%) covering 23% of atlas.pPaHC r (Parahippocampal Gyrus, posterior division Right)
56 voxels (5%) covering 14% of atlas.pPaHC l (Parahippocampal Gyrus, posterior division Left)
49 voxels (4%) covering 15% of atlas.Amygdala l
44 voxels (4%) covering 19% of atlas.Ver3 (Vermis 3)
34 voxels (3%) covering 4% of atlas.Cereb45 l (Cerebelum 4 5 Left)
25 voxels (2%) covering 8% of atlas.aTFusC l (Temporal Fusiform Cortex, anterior division Left)
19 voxels (2%) covering 3% of atlas.Ver45 (Vermis 4 5)
16 voxels (1%) covering 0% of atlas.Brain-Stem
12 voxels (1%) covering 1% of atlas.FOrb l (Frontal Orbital Cortex Left)
11 voxels (1%) covering 2% of atlas.Cereb45 r (Cerebelum 4 5 Right)
10 voxels (1%) covering 1% of atlas.LG r (Lingual Gyrus Right)
7 voxels (1%) covering 4% of atlas.Cereb3 r (Cerebelum 3 Right)
5 voxels (0%) covering 1% of atlas.Hippocampus r
339 voxels (29%) covering 0% of atlas.not-labeled

**Table S26. SBC map association with MCT using the whole Precuneus as seed.**

| Cluster (x,y,z) | Size (vox) | size p-FWE | size p-FDR | size p-unc | peak p-FWE | peak p-unc |
|---|---|---|---|---|---|---|
| +00 -52 +58 | 4088 | 0.000000 | 0.000000 | 0.000000 | 0.784095 | 0.000023 |
| +16 -34 -12 | 979 | 0.056464 | 0.022278 | 0.000199 | 0.804811 | 0.000025 |
| +60 -62 -18 | 949 | 0.066894 | 0.022278 | 0.000237 | 0.990956 | 0.000092 |

**Cluster +0 -52 +58**:
2314 voxels (57%) covering 41% of atlas.Precuneous (Precuneous Cortex)
414 voxels (10%) covering 17% of atlas.PC (Cingulate Gyrus, posterior division)
141 voxels (3%) covering 10% of atlas.SPL l (Superior Parietal Lobule Left)
101 voxels (2%) covering 16% of atlas.Cuneal r (Cuneal Cortex Right)
64 voxels (2%) covering 2% of atlas.AC (Cingulate Gyrus, anterior division)
48 voxels (1%) covering 1% of atlas.sLOC r (Lateral Occipital Cortex, superior division Right)
40 voxels (1%) covering 8% of atlas.Cuneal l (Cuneal Cortex Left)

32 voxels (1%) covering 5% of atlas.SMA L(Juxtapositional Lobule Cortex -formerly Supplementary Motor Cortex- Left)
28 voxels (1%) covering 1% of atlas.PreCG l (Precentral Gyrus Left)
24 voxels (1%) covering 1% of atlas.PostCG l (Postcentral Gyrus Left)
15 voxels (0%) covering 0% of atlas.PreCG r (Precentral Gyrus Right)
5 voxels (0%) covering 3% of atlas.SCC r (Supracalcarine Cortex Right)
4 voxels (0%) covering 0% of atlas.PostCG r (Postcentral Gyrus Right)
2 voxels (0%) covering 0% of atlas.sLOC l (Lateral Occipital Cortex, superior division Left)
2 voxels (0%) covering 0% of atlas.PaCiG l (Paracingulate Gyrus Left)
854 voxels (21%) covering 0% of atlas.not-labeled

**Cluster +16 -34 -12**:
150 voxels (15%) covering 47% of atlas.pPaHC r (Parahippocampal Gyrus, posterior division Right)
98 voxels (10%) covering 25% of atlas.pPaHC l (Parahippocampal Gyrus, posterior division Left)
64 voxels (7%) covering 7% of atlas.Cereb45 l (Cerebelum 4 5 Left)
64 voxels (7%) covering 10% of atlas.Ver45 (Vermis 4 5)
63 voxels (6%) covering 9% of atlas.Hippocampus r
61 voxels (6%) covering 27% of atlas.Ver3 (Vermis 3)
48 voxels (5%) covering 7% of atlas.aPaHC r (Parahippocampal Gyrus, anterior division Right)
40 voxels (4%) covering 1% of atlas.Brain-Stem
35 voxels (4%) covering 4% of atlas.TOFusC r (Temporal Occipital Fusiform Cortex Right)
33 voxels (3%) covering 2% of atlas.LG r (Lingual Gyrus Right)
33 voxels (3%) covering 5% of atlas.Cereb45 r (Cerebelum 4 5 Right)
29 voxels (3%) covering 16% of atlas.Cereb3 r (Cerebelum 3 Right)
15 voxels (2%) covering 2% of atlas.Hippocampus l
11 voxels (1%) covering 8% of atlas.Cereb3 l (Cerebelum 3 Left)
6 voxels (1%) covering 1% of atlas.pTFusC r (Temporal Fusiform Cortex, posterior division Right)
3 voxels (0%) covering 0% of atlas.LG l (Lingual Gyrus Left)
226 voxels (23%) covering 0% of atlas.not-labeled

**Cluster +60 -62 -18**:
275 voxels (29%) covering 22% of atlas.pSMG r (Supramarginal Gyrus, posterior division Right)
116 voxels (12%) covering 8% of atlas.AG r (Angular Gyrus Right)
80 voxels (8%) covering 6% of atlas.pMTG r (Middle Temporal Gyrus, posterior division Right)
72 voxels (8%) covering 9% of atlas.toITG r (Inferior Temporal Gyrus, temporooccipital part Right)
60 voxels (6%) covering 14% of atlas.pSTG r (Superior Temporal Gyrus, posterior division Right)
53 voxels (6%) covering 12% of atlas.PT r (Planum Temporale Right)
35 voxels (4%) covering 3% of atlas.toMTG r (Middle Temporal Gyrus, temporooccipital part Right)
18 voxels (2%) covering 3% of atlas.PO r (Parietal Operculum Cortex Right)
16 voxels (2%) covering 2% of atlas.aSMG r (Supramarginal Gyrus, anterior division Right)
224 voxels (24%) covering 0% of atlas.not-labeled

**Table S27. SBC map association with MCT using 7Pm as seed.**

| Cluster (x,y,z) | Size (vox) | size p-FWE | size p-FDR | size p-unc | peak p-FWE | peak p-unc |
|---|---|---|---|---|---|---|
| -30 -40 -08 | 1046 | 0.041906 | 0.043850 | 0.000149 | 0.885375 | 0.000035 |

**Cluster -30 -40 -8**:
187 voxels (18%) covering 3% of atlas.Precuneous (Precuneous Cortex)

161 voxels (15%) covering 21% of atlas.Hippocampus l
157 voxels (15%) covering 40% of atlas.pPaHC l (Parahippocampal Gyrus, posterior division Left)
86 voxels (8%) covering 6% of atlas.LG l (Lingual Gyrus Left)
48 voxels (5%) covering 6% of atlas.pTFusC l (Temporal Fusiform Cortex, posterior division Left)
39 voxels (4%) covering 1% of atlas.Brain-Stem
22 voxels (2%) covering 1% of atlas.PC (Cingulate Gyrus, posterior division)
14 voxels (1%) covering 2% of atlas.Cereb45 l (Cerebelum 4 5 Left)
10 voxels (1%) covering 2% of atlas.Ver45 (Vermis 4 5)
8 voxels (1%) covering 1% of atlas.aPaHC l (Parahippocampal Gyrus, anterior division Left)
6 voxels (1%) covering 1% of atlas.TOFusC l (Temporal Occipital Fusiform Cortex Left)
5 voxels (0%) covering 1% of atlas.ICC l (Intracalcarine Cortex Left)
5 voxels (0%) covering 4% of atlas.Cereb3 l (Cerebelum 3 Left)
3 voxels (0%) covering 2% of atlas.SCC r (Supracalcarine Cortex Right)
2 voxels (0%) covering 0% of atlas.Thalamus l
293 voxels (28%) covering 0% of atlas.not-labeled

**Table S28. SBC map association with MCT using 7Am as seed.**

| Cluster (x,y,z) | Size (vox) | size p-FWE | size p-FDR | size p-unc | peak p-FWE | peak p-unc |
|---|---|---|---|---|---|---|
| +06 -90 +18 | 2943 | 0.000007 | 0.000007 | 0.000000 | 0.788972 | 0.000024 |

**Cluster +6 -90 +18**:
1269 voxels (43%) covering 23% of atlas.Precuneous (Precuneous Cortex)
403 voxels (14%) covering 8% of atlas.sLOC l (Lateral Occipital Cortex, superior division Left)
319 voxels (11%) covering 42% of atlas.ICC r (Intracalcarine Cortex Right)
135 voxels (5%) covering 21% of atlas.ICC l (Intracalcarine Cortex Left)
112 voxels (4%) covering 22% of atlas.Cuneal l (Cuneal Cortex Left)
108 voxels (4%) covering 5% of atlas.PC (Cingulate Gyrus, posterior division)
60 voxels (2%) covering 9% of atlas.Cuneal r (Cuneal Cortex Right)
60 voxels (2%) covering 3% of atlas.LG r (Lingual Gyrus Right)
55 voxels (2%) covering 38% of atlas.SCC r (Supracalcarine Cortex Right)
50 voxels (2%) covering 3% of atlas.LG l (Lingual Gyrus Left)
48 voxels (2%) covering 3% of atlas.SPL l (Superior Parietal Lobule Left)
35 voxels (1%) covering 1% of atlas.OP r (Occipital Pole Right)
19 voxels (1%) covering 0% of atlas.sLOC r (Lateral Occipital Cortex, superior division Right)
13 voxels (0%) covering 18% of atlas.SCC l (Supracalcarine Cortex Left)
11 voxels (0%) covering 0% of atlas.PostCG l (Postcentral Gyrus Left)
2 voxels (0%) covering 0% of atlas.Ver45 (Vermis 4 5)
1 voxels (0%) covering 0% of atlas.Cereb45 l (Cerebelum 4 5 Left)
243 voxels (8%) covering 0% of atlas.not-labeled

**Table S29. SBC map association with MCT using 7m as seed.**

| Cluster (x,y,z) | Size (vox) | size p-FWE | size p-FDR | size p-unc | peak p-FWE | peak p-unc |
|---|---|---|---|---|---|---|

| | | | | | | |
|---|---|---|---|---|---|---|
| +60 -36 +20 | 3859 | 0.000000 | 0.000000 | 0.000000 | 0.299732 | 0.000004 |

**Cluster +60 -36 +20**:
1398 voxels (36%) covering 25% of atlas.Precuneous (Precuneous Cortex)
516 voxels (13%) covering 22% of atlas.PC (Cingulate Gyrus, posterior division)
470 voxels (12%) covering 32% of atlas.AG r (Angular Gyrus Right)
342 voxels (9%) covering 7% of atlas.sLOC r (Lateral Occipital Cortex, superior division Right)
193 voxels (5%) covering 16% of atlas.pSMG r (Supramarginal Gyrus, posterior division Right)
73 voxels (2%) covering 11% of atlas.Cuneal r (Cuneal Cortex Right)
61 voxels (2%) covering 2% of atlas.AC (Cingulate Gyrus, anterior division)
46 voxels (1%) covering 10% of atlas.PT r (Planum Temporale Right)
31 voxels (1%) covering 2% of atlas.SPL r (Superior Parietal Lobule Right)
22 voxels (1%) covering 2% of atlas.SPL l (Superior Parietal Lobule Left)
21 voxels (1%) covering 4% of atlas.PO r (Parietal Operculum Cortex Right)
19 voxels (0%) covering 1% of atlas.PostCG l (Postcentral Gyrus Left)
17 voxels (0%) covering 0% of atlas.PreCG r (Precentral Gyrus Right)
13 voxels (0%) covering 0% of atlas.PreCG l (Precentral Gyrus Left)
9 voxels (0%) covering 1% of atlas.SMA r (Juxtapositional Lobule Cortex -formerly Supplementary Motor Cortex- Right)
3 voxels (0%) covering 0% of atlas.PostCG r (Postcentral Gyrus Right)
1 voxels (0%) covering 0% of atlas.sLOC l (Lateral Occipital Cortex, superior division Left)
624 voxels (16%) covering 0% of atlas.not-labeled

**Table S30. SBC map association with MCT using PCV as seed.**

| Cluster (x,y,z) | Size (vox) | size p-FWE | size p-FDR | size p-unc | peak p-FWE | peak p-unc |
|---|---|---|---|---|---|---|
| -02 -50 +56 | 2656 | 0.000023 | 0.000022 | 0.000000 | 0.509630 | 0.000009 |
| +02 -36 -40 | 2355 | 0.000081 | 0.000040 | 0.000000 | 0.981858 | 0.000077 |
| +08 +52 -18 | 1216 | 0.017339 | 0.005681 | 0.000061 | 0.999317 | 0.000164 |

**Cluster -2 -50 +56**:
1612 voxels (61%) covering 29% of atlas.Precuneous (Precuneous Cortex)
144 voxels (5%) covering 6% of atlas.PC (Cingulate Gyrus, posterior division)
100 voxels (4%) covering 13% of atlas.ICC r (Intracalcarine Cortex Right)
95 voxels (4%) covering 7% of atlas.SPL l (Superior Parietal Lobule Left)
48 voxels (2%) covering 7% of atlas.Cuneal r (Cuneal Cortex Right)
44 voxels (2%) covering 31% of atlas.SCC r (Supracalcarine Cortex Right)
38 voxels (1%) covering 6% of atlas.ICC l (Intracalcarine Cortex Left)
31 voxels (1%) covering 1% of atlas.PostCG l (Postcentral Gyrus Left)
20 voxels (1%) covering 4% of atlas.Cuneal l (Cuneal Cortex Left)
20 voxels (1%) covering 1% of atlas.Thalamus l
18 voxels (1%) covering 3% of atlas.Ver45 (Vermis 4 5)
11 voxels (0%) covering 0% of atlas.Brain-Stem
5 voxels (0%) covering 7% of atlas.SCC l (Supracalcarine Cortex Left)

4 voxels (0%) covering 1% of atlas.Hippocampus l
2 voxels (0%) covering 0% of atlas.sLOC r (Lateral Occipital Cortex, superior division Right)
2 voxels (0%) covering 0% of atlas.LG r (Lingual Gyrus Right)
2 voxels (0%) covering 0% of atlas.LG l (Lingual Gyrus Left)
2 voxels (0%) covering 0% of atlas.Cereb45 l (Cerebelum 4 5 Left)
1 voxels (0%) covering 0% of atlas.SPL r (Superior Parietal Lobule Right)
457 voxels (17%) covering 0% of atlas.not-labeled

**Cluster +2 -36 -40**:
482 voxels (20%) covering 21% of atlas.Cereb8 r (Cerebelum 8 Right)
440 voxels (19%) covering 17% of atlas.Cereb1 r (Cerebelum Crus1 Right)
309 voxels (13%) covering 7% of atlas.Brain-Stem
157 voxels (7%) covering 7% of atlas.Cereb2 r (Cerebelum Crus2 Right)
89 voxels (4%) covering 37% of atlas.Ver8 (Vermis 8)
71 voxels (3%) covering 5% of atlas.Cereb6 r (Cerebelum 6 Right)
57 voxels (2%) covering 11% of atlas.Cereb7 r (Cerebelum 7b Right)
54 voxels (2%) covering 3% of atlas.Cereb8 l (Cerebelum 8 Left)
31 voxels (1%) covering 21% of atlas.Cereb10 l (Cerebelum 10 Left)
22 voxels (1%) covering 3% of atlas.aPaHC r (Parahippocampal Gyrus, anterior division Right)
22 voxels (1%) covering 3% of atlas.Cereb9 l (Cerebelum 9 Left)
21 voxels (1%) covering 13% of atlas.Cereb10 r (Cerebelum 10 Right)
20 voxels (1%) covering 12% of atlas.Ver9 (Vermis 9)
16 voxels (1%) covering 3% of atlas.aPaHC l (Parahippocampal Gyrus, anterior division Left)
14 voxels (1%) covering 2% of atlas.Cereb45 r (Cerebelum 4 5 Right)
12 voxels (1%) covering 2% of atlas.Cereb9 r (Cerebelum 9 Right)
12 voxels (1%) covering 6% of atlas.Ver7 (Vermis 7)
8 voxels (0%) covering 1% of atlas.Cereb7 l (Cerebelum 7b Left)
5 voxels (0%) covering 0% of atlas.Cereb2 l (Cerebelum Crus2 Left)
4 voxels (0%) covering 4% of atlas.Ver10 (Vermis 10)
1 voxels (0%) covering 0% of atlas.pTFusC r (Temporal Fusiform Cortex, posterior division Right)
508 voxels (22%) covering 0% of atlas.not-labeled

**Cluster +8 +52 -18**:
363 voxels (30%) covering 37% of atlas.MedFC (Frontal Medial Cortex)
221 voxels (18%) covering 20% of atlas.SubCalC (Subcallosal Cortex)
116 voxels (10%) covering 2% of atlas.FP l (Frontal Pole Left)
104 voxels (9%) covering 1% of atlas.FP r (Frontal Pole Right)
24 voxels (2%) covering 2% of atlas.PaCiG l (Paracingulate Gyrus Left)
11 voxels (1%) covering 1% of atlas.PaCiG r (Paracingulate Gyrus Right)
6 voxels (0%) covering 0% of atlas.FOrb r (Frontal Orbital Cortex Right)
5 voxels (0%) covering 0% of atlas.FOrb l (Frontal Orbital Cortex Left)
2 voxels (0%) covering 0% of atlas.AC (Cingulate Gyrus, anterior division)
364 voxels (30%) covering 0% of atlas.not-labeled

**Table S31. SBC map association with MCT using POS2 as seed.**

| Cluster (x,y,z) | Size (vox) | size p-FWE | size p-FDR | size p-unc | peak p-FWE | peak p-unc |
|---|---|---|---|---|---|---|
| -02 -50 +56 | 2656 | 0.000023 | 0.000022 | 0.000000 | 0.509630 | 0.000009 |
| +02 -36 -40 | 2355 | 0.000081 | 0.000040 | 0.000000 | 0.981858 | 0.000077 |
| +08 +52 -18 | 1216 | 0.017339 | 0.005681 | 0.000061 | 0.999317 | 0.000164 |

**Cluster -2 -50 +56**:
1612 voxels (61%) covering 29% of atlas.Precuneous (Precuneous Cortex)
144 voxels (5%) covering 6% of atlas.PC (Cingulate Gyrus, posterior division)
100 voxels (4%) covering 13% of atlas.ICC r (Intracalcarine Cortex Right)
95 voxels (4%) covering 7% of atlas.SPL l (Superior Parietal Lobule Left)
48 voxels (2%) covering 7% of atlas.Cuneal r (Cuneal Cortex Right)
44 voxels (2%) covering 31% of atlas.SCC r (Supracalcarine Cortex Right)
38 voxels (1%) covering 6% of atlas.ICC l (Intracalcarine Cortex Left)
31 voxels (1%) covering 1% of atlas.PostCG l (Postcentral Gyrus Left)
20 voxels (1%) covering 4% of atlas.Cuneal l (Cuneal Cortex Left)
20 voxels (1%) covering 1% of atlas.Thalamus l
18 voxels (1%) covering 3% of atlas.Ver45 (Vermis 4 5)
11 voxels (0%) covering 0% of atlas.Brain-Stem
5 voxels (0%) covering 7% of atlas.SCC l (Supracalcarine Cortex Left)
4 voxels (0%) covering 1% of atlas.Hippocampus l
2 voxels (0%) covering 0% of atlas.sLOC r (Lateral Occipital Cortex, superior division Right)
2 voxels (0%) covering 0% of atlas.LG r (Lingual Gyrus Right)
2 voxels (0%) covering 0% of atlas.LG l (Lingual Gyrus Left)
2 voxels (0%) covering 0% of atlas.Cereb45 l (Cerebelum 4 5 Left)
1 voxels (0%) covering 0% of atlas.SPL r (Superior Parietal Lobule Right)
457 voxels (17%) covering 0% of atlas.not-labeled

**Cluster +2 -36 -40**:
482 voxels (20%) covering 21% of atlas.Cereb8 r (Cerebelum 8 Right)
440 voxels (19%) covering 17% of atlas.Cereb1 r (Cerebelum Crus1 Right)
309 voxels (13%) covering 7% of atlas.Brain-Stem
157 voxels (7%) covering 7% of atlas.Cereb2 r (Cerebelum Crus2 Right)
89 voxels (4%) covering 37% of atlas.Ver8 (Vermis 8)
71 voxels (3%) covering 5% of atlas.Cereb6 r (Cerebelum 6 Right)
57 voxels (2%) covering 11% of atlas.Cereb7 r (Cerebelum 7b Right)
54 voxels (2%) covering 3% of atlas.Cereb8 l (Cerebelum 8 Left)
31 voxels (1%) covering 21% of atlas.Cereb10 l (Cerebelum 10 Left)
22 voxels (1%) covering 3% of atlas.aPaHC r (Parahippocampal Gyrus, anterior division Right)
22 voxels (1%) covering 3% of atlas.Cereb9 l (Cerebelum 9 Left)
21 voxels (1%) covering 13% of atlas.Cereb10 r (Cerebelum 10 Right)
20 voxels (1%) covering 12% of atlas.Ver9 (Vermis 9)
16 voxels (1%) covering 3% of atlas.aPaHC l (Parahippocampal Gyrus, anterior division Left)

14 voxels (1%) covering 2% of atlas.Cereb45 r (Cerebelum 4 5 Right)
12 voxels (1%) covering 2% of atlas.Cereb9 r (Cerebelum 9 Right)
12 voxels (1%) covering 6% of atlas.Ver7 (Vermis 7)
8 voxels (0%) covering 1% of atlas.Cereb7 l (Cerebelum 7b Left)
5 voxels (0%) covering 0% of atlas.Cereb2 l (Cerebelum Crus2 Left)
4 voxels (0%) covering 4% of atlas.Ver10 (Vermis 10)
1 voxels (0%) covering 0% of atlas.pTFusC r (Temporal Fusiform Cortex, posterior division Right)
508 voxels (22%) covering 0% of atlas.not-labeled

**Cluster +8 +52 -18**:
363 voxels (30%) covering 37% of atlas.MedFC (Frontal Medial Cortex)
221 voxels (18%) covering 20% of atlas.SubCalC (Subcallosal Cortex)
116 voxels (10%) covering 2% of atlas.FP l (Frontal Pole Left)
104 voxels (9%) covering 1% of atlas.FP r (Frontal Pole Right)
24 voxels (2%) covering 2% of atlas.PaCiG l (Paracingulate Gyrus Left)
11 voxels (1%) covering 1% of atlas.PaCiG r (Paracingulate Gyrus Right)
6 voxels (0%) covering 0% of atlas.FOrb r (Frontal Orbital Cortex Right)
5 voxels (0%) covering 0% of atlas.FOrb l (Frontal Orbital Cortex Left)
2 voxels (0%) covering 0% of atlas.AC (Cingulate Gyrus, anterior division)
364 voxels (30%) covering 0% of atlas.not-labeled
